# Online monitoring technology for deep phenotyping of cognitive impairment after stroke

**DOI:** 10.1101/2024.09.06.24313173

**Authors:** Dragos-Cristian Gruia, Valentina Giunchiglia, Aoife Coghlan, Sophie Brook, Soma Banerjee, Jo Kwan, Peter J. Hellyer, Adam Hampshire, Fatemeh Geranmayeh

## Abstract

**Background:** Despite the high prevalence of disabling post-stroke cognitive sequalae, these impairments are often underdiagnosed and rarely monitored longitudinally. Provision of unsupervised remote online cognitive technology would provide a scalable solution to this problem. However, despite recent advances, such technology is currently lacking, with existing tools either not meeting the scalability challenge or not optimised for specific applications in post-stroke cognitive impairment. To address this gap, we designed and developed a comprehensive online battery highly optimised for detecting cognitive impairments in stroke survivors.

**Method:** The technology is optimised to allow both diagnosis and monitoring of post-stroke deficits, and for remote unsupervised administration. Participants performed 22 computerised tasks, and answered neuropsychiatric questionnaires and patient reported outcomes. 90 stroke survivors (Mean age = 62.1 years; 68% and 32% in the acute and subacute/chronic phase after stroke respectively) and over 6,000 age-matched healthy older adults were recruited. Patient outcome measures were derived from Bayesian Regression modelling of the large normative sample and validated against standard clinical scales.

**Results:** Our online technology has greater sensitivity to post-stroke cognitive impairment than pen-and-paper tests such as the MOCA (mean sensitivity 81.75% and 52.25% respectively, P<0.001). Further, our outcomes show a stronger correlation with post-stroke quality of life (r(78)=0.51, R2=0.26, P<0.001) when compared to MOCA, which only explains half of this variance (r(78)=0.38, R2=0.14, P< 0.001). An additional set of experiments confirm that the online tasks yield highly reliable outcomes, with consistent performance observed across supervised versus unsupervised settings, and minimal learning effects across multiple timepoints.

**Conclusion:** The current online cognitive monitoring technology is feasible, sensitive, and reliable when assessing patients with stroke. The technology offers an economical and scalable method for assessing post-stroke cognition in the clinical setting and sensitively monitoring cognitive outcomes in clinical trials for stroke.

## Introduction

Stroke is a leading cause of death and disability globally, with cognitive sequalae affecting three-quarters of survivors.^1^ The spectrum of cognitive impairments associated with stroke encompasses domain-specific deficits, such as aphasia, neglect, and memory impairment as well as domain-general deficits usually associated with co-existing small vessel disease such as executive / attentional dysfunction and reduction in processing speed.^2^ Collectively, these impairments have a detrimental impact on poststroke recovery, engagement with therapeutic interventions and lower quality of life among patients.^3,4^ Consequently, early detection of these impairments has been recommended by key stake holders and national and international guidelines for stroke management.^5–7^

Despite a lack of universally accepted approaches for identifying post-stroke cognitive deficits, there is consensus that stroke survivors should be screened and monitored for cognitive impairment using a stroke-specific cognitive assessment.^5,8^ There remains considerable variability in clinical practice, with deployed assessments ranging from extensive neuropsychological test batteries tailored to a specific cognitive domain (e.g., language), to global measures of cognition using brief screening tools like the MoCA which were not developed for stroke.^9^ This choice is often driven by personal preferences, availability, cost and time pressures, leaving little prospect for generalisability of findings across sites.

Availability of a cost-effective, reliable, scalable and comprehensive screening tool that provides a stroke-specific deep phenotyping of cognition would be transformative for clinical diagnosis, as well as enabling much-needed large-scale population-based research for studying the mechanisms of post-stroke cognitive recovery. To address this gap, we developed a novel digital adaptive technology: The Imperial Comprehensive Cognitive Assessment in Cerebrovascular Disease (IC3).^10^ IC3 is a digital assessment battery highly optimised for stroke survivors and designed to require minimal input from a clinician in detecting both domain-general and domain-specific cognitive deficits in patients after stroke. It provides a comprehensive profile of performance across cognitive domains known to be impaired following stroke, including memory, language, executive function, attention, numeracy, praxis as well as hand motor ability and clinical and neuropsychiatric questionnaires.

First, we present and analyse extensive normative data derived from over 6,000 UK-based older adults using the IC3 technology, highlighting its ability to map cognition at a large scale, in a time- and cost-efficient manner. Leveraging the large sample size, and state-of-the-art Bayesian modelling, we create patient-specific predictive scores that account for the effects of demographic and neuropsychiatric variables as well as language proficiency, dyslexia and device on cognitive performance. IC3’s validity as a remote cognitive screening tool is tested through a robust set of sub-analyses that quantify its reliability and feasibility, equivalence in performance between supervised and non-supervised settings, and learning effects across 4 timepoints.

We examine whether in patients with stroke, the IC3-derived scores map onto well-established first-line clinical screening tools (MoCA), have comparable sensitivity to mild cognitive impairment and with patient reported functional outcome measures (post-stroke quality of life), and explain as much or more variation than the first-line clinical screening tool (MoCA). Using factor analysis, we examine whether IC3 scores map intuitively onto cognitive domains often affected in stroke. We discuss the results in relation to the feasibility and validity of the IC3 battery as a technology for monitoring cognition across populations with stroke and cerebrovascular diseases.

## Method

### Participants

The different participant cohorts employed in this study are described below and shown in Figure 1.

**Figure 1.**
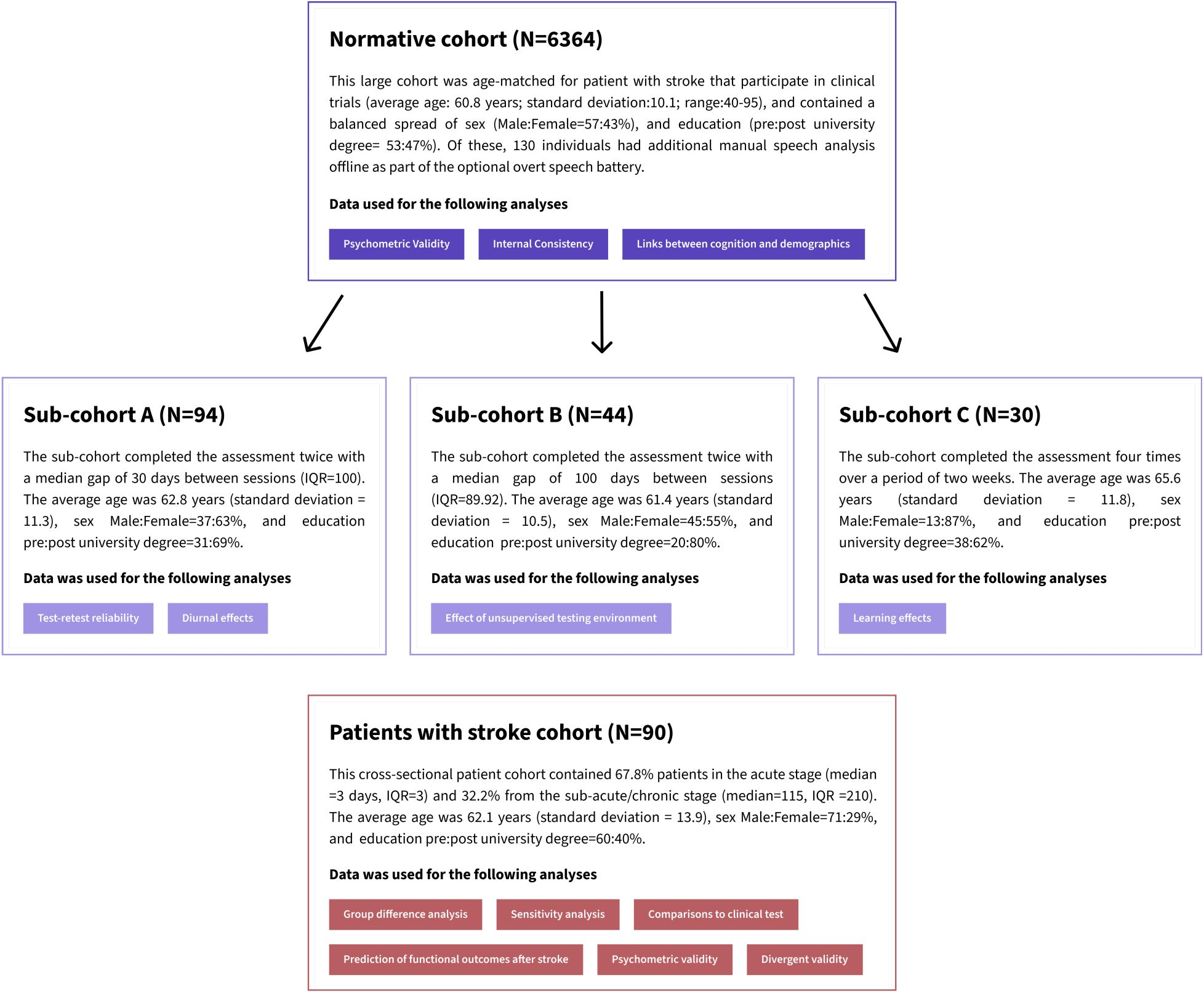
Overview of the study cohorts discussed in this study, along with the analysis conducted on each group.

#### Normative cohort

A study invitation was extended to 25,000 individuals, over the age of 40, residing in Great Britain, all of whom had previously participated in the Great British Intelligence Test (a nationwide initiative aimed at mapping cognition within the general population) and consented to being recontacted for other studies.^11^ Ultimately, 7,095 healthy older adults provided their consent and initiated the IC3 cognitive battery, with 5,639 participants (79.5%) successfully completing all 22 tasks. The data were collected remotely online, between October and November 2022. Participation in the study was voluntary with no monetary incentive. In addition, we collected data from 138 individuals via the Imperial Clinical Research Facility participant registry for reliability and validity purposes. The combined normative cohort contained 6364 healthy older adults, following participant exclusion and pre-processing steps (detailed in Supplementary Material 3).

#### Normative sub-cohorts used for the reliability and validity analyses (**Figure 1**, Sub Cohort A-C)

A smaller sample of controls who performed the assessment multiple times was collected separately to test the battery’s reliability and validity. These were recruited using the Imperial Clinical Research Facility participant registry. A total of 94 participants (Sub-cohort A) completed the assessment twice, as part of the test-retest reliability analysis. Of the 94 participants, 44 (Sub-cohort B) performed the assessment both supervised (in person), and unsupervised (remote) with the order counterbalanced. The remainder (50/94) completed both sessions unsupervised. A subset (N=30/94; Sub-Cohort C) completed the assessment remotely 4 times in total over the course of two weeks, to allow for the estimation of any potential learning effects. No monetary reward was provided, aside from travel reimbursements where appropriate.

#### Patients with stroke cohort

Patients with radiologically confirmed stroke were recruited from Imperial College Healthcare NHS Trust. Exclusion criteria included pre-stroke diagnosis of dementia, pure brain stem stroke, severe visuospatial problems, severe mental health diagnoses, fatigue limiting engagement with the IC3 beyond 15 minutes and inability to understand task instructions. Consecutively recruited patients underwent the digital IC3 assessment (see Supplementary Material 5.1. for detailed demographic information). The scores were compared with clinical pen-and-paper cognitive screens (MoCA).

All participants gave informed consent. The data was acquired as part of a longitudinal observational clinical study approved by UK’s Health Research Authority (Registered under NCT05885295; IRAS:299333; REC:21/SW/0124). Patients also underwent blood biomarker testing and brain imaging which will not be analysed in this paper. A lesion overlap map is shown in Supplementary Material 5.2. demonstrates the lesion distribution in patients who had MRI brain imaging.

### Cognitive tasks, speech-based tasks and neuropsychiatric questionnaires

A graphical overview and a detailed description of the cognitive assessments are available in Figure 2 and Supplementary Material 1. respectively. These cover 18 short cognitive tasks with additional 4 optional speech production tasks, collectively covering a wide range of cognitive domains known to be affected post-stroke. The tasks were followed by clinically-validated questionnaires (Apathy Evaluation Scale Fatigue Scale, Geriatric Depression Scale, and Instrumental Activities of Daily Living).^12–15^

**Figure 2.**
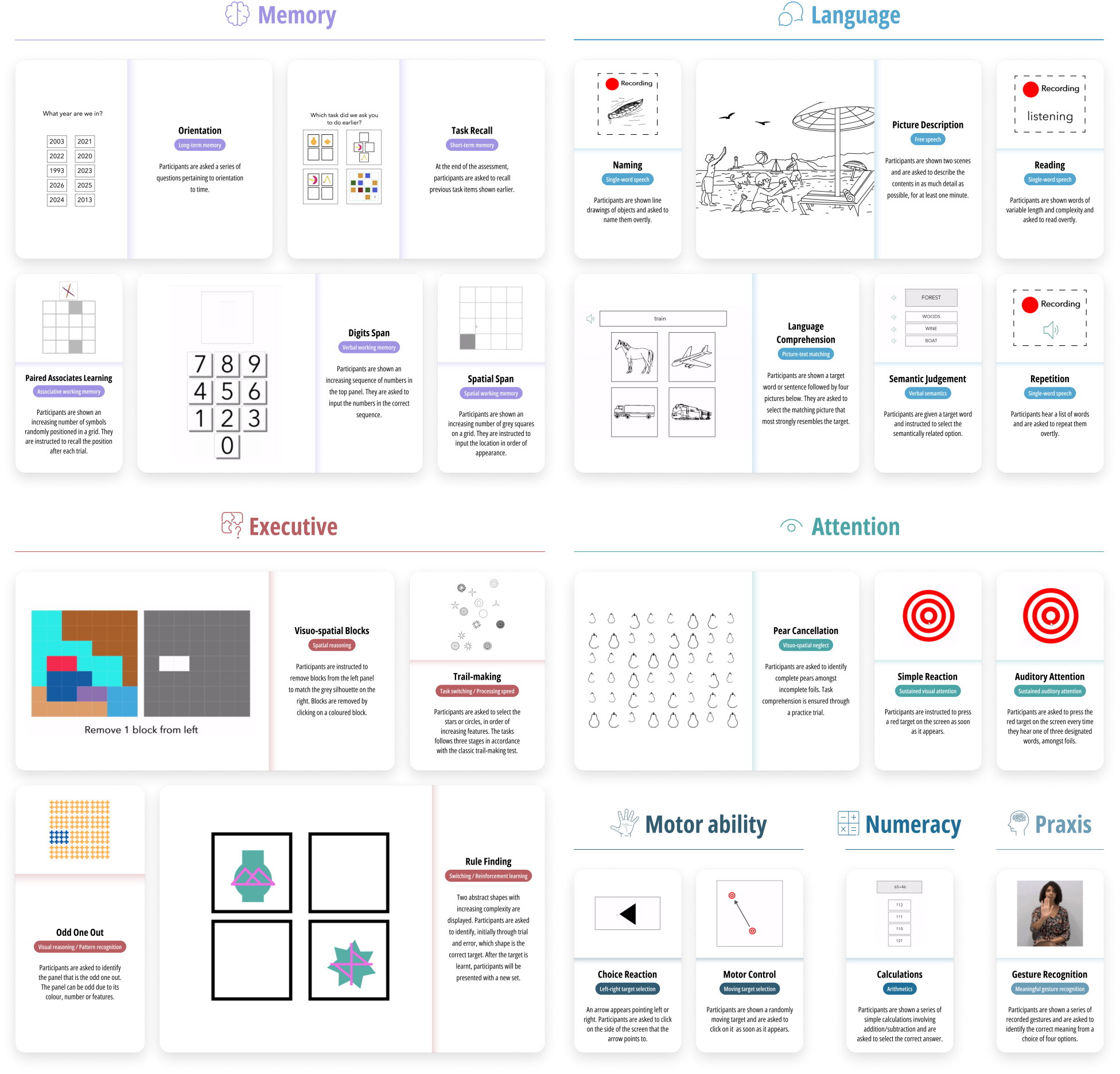
Graphical overview of 22 IC3 tasks organised by the main cognitive domains tested: memory, language, executive, attention, motor ability, numeracy and praxis. The four optional speech production tasks (naming, repetition, reading, picture description) allow speech to be recorded and manually analysed offline.

IC3 is deployable via a web-browser on practically any modern smartphone, tablet, or computer/laptop device via a weblink and is implemented through Cognitron (https://www.cognitron.co.uk/), a state-of-the art platform for remote neuropsychological testing that is rapidly being adopted by large scale population studies both in the UK and internationally.^16,17^

### Cognitive and speech data pre-processing

Given the remote nature of the cognitive testing, to ensure that the normative data was derived from fully engaged, healthy participants who understood the task instructions, we implemented three levels of data filtering (subject-level, task-level, and trial-level), by filtering out data that was invalid or of poor quality. See Supplementary Materials 3. for a detailed description. For the four optional speech production tasks data was manually analysed offline by 6 trained expert raters with high inter-rater reliability (average intraclass correlation=0.86). See Supplementary Materials 2 for speech marking guidelines.

### IC3 validation

A series of analyses quantified IC3’s reliability, equivalent performance between supervised and non-supervised settings, and learning effects (see Supplementary Materials 5.3. – 5.7. for more details).

### Bayesian modelling on the large normative sample derives patient-specific predictive scores

Constrained by relatively small normative sample sizes, existing cognitive batteries have traditionally been limited to accounting for the effects of demographic factors by stratifying the normative sample into even smaller sub-groups, often limited to one or two variables (age and occasionally education). In this study, we leverage the large normative sample of 6364 individuals and Bayesian modelling (see below), to create patient-specific predictive scores with higher precision, accounting for 8 additional confounding factors (age, sex, education, language proficiency, testing device, depression, dyslexia, anxiety; highlighted with an asterisk in Supplementary Material 5.1.).

#### State-of-the-art Bayesian posterior predictions for modelling the relationship between cognition and confounding factors in controls

Bayesian regression analyses containing all 8 covariates were performed separately for all tasks, to estimate the effects of each of the covariates on individual task performance. For the optional speech production tasks (repetition, naming, reading), the smaller sample (N=130) precluded the inclusion of depression, anxiety and dyslexia as covariates. For full details of Bayesian regression models and how the coefficient were derived, see Supplementary Materials 4.1.-4.2.).

#### Patient-specific impairment thresholds

Bayesian modelling was used to create patient-specific impairment thresholds correcting for the aforementioned confounding variables. This was done by (i) training the Bayesian regression models on the normative sample as described above, (ii) using the derived posterior distributions to estimate patient-specific predicted performance, converted to standard deviation (SD) units, and iii) subtracting the observed patient performance (also in SD units) from the predicted performance derived from step ‘ii’. A resulting negative “deviation from norm” score suggests that the patient had a deficit in that specific task by a given magnitude in SD units, such that a score of -1 represents an impairment of 1 SD from a corresponding demographically matched control group. Using these estimates, boundaries for the severity of the cognitive impairments were arbitrarily assigned as -1.5 (mild), -2.0 (moderate) and -2.5 (severe) SD below the mean, in line with previous post-stroke cognitive tests.^18^

## Results

### Normative sample

#### Participant characteristics

The cleaned normative sample consisted of 6364 individuals. The number of participants were varied for each task (N= 4782-6290) depending on task-specific data filtering and the fact that tasks at the end of the assessment had fewer timepoints. Supplementary Material 5.1. outlines the demographics and additional confounding factors that may affect cognitive performance.

#### Relationships between cognition and confounding demographic factors in the normative sample

Standardised coefficients were obtained from task-specific Bayesian regression models for the 8 confounding covariates (mean R^2^:11.29%; range: 1.3-53.5). The lower range of the R^2^ was driven by tasks with ceiling effects and low inter-subject variability in the controls (e.g., 3.9/4.0 and 3.8/4.00 for Orientation and Task Recall mean performance respectively).

The strength of the association between each of the 8 covariates and cognition are shown in Figure 3 where warm and cool colours represent positive and negative standardised coefficients. Cognitive performance generally worsened with age as shown by negative coefficients shown in purple. The exception is Semantic Judgement in keeping with previous literature demonstrating age-related improvement in language function.^19^ Dyslexia and English as a second language had a strong negative effect on performance particularly on tasks involving language and numeracy skills. Device was a strong confounding factor in task that relied on speed and motor dexterity as the main outcome measure (e.g. Simple Rection Time, Choice Reaction Time and Motor Control). This association is understandable given faster responses on touch screen compared to mouse/trackpad-operated devices. Higher education levels were related to better cognitive performance across tasks that involved language and numeracy, with the least effect on tasks that primarily captured motor dexterity (e.g., Motor Control, Choice Reaction Task, Simple Reaction Time tasks). Overall, the regression models provide intuitive and interpretable relationships between cognition and the 8 confounding factors.

**Figure 3.**
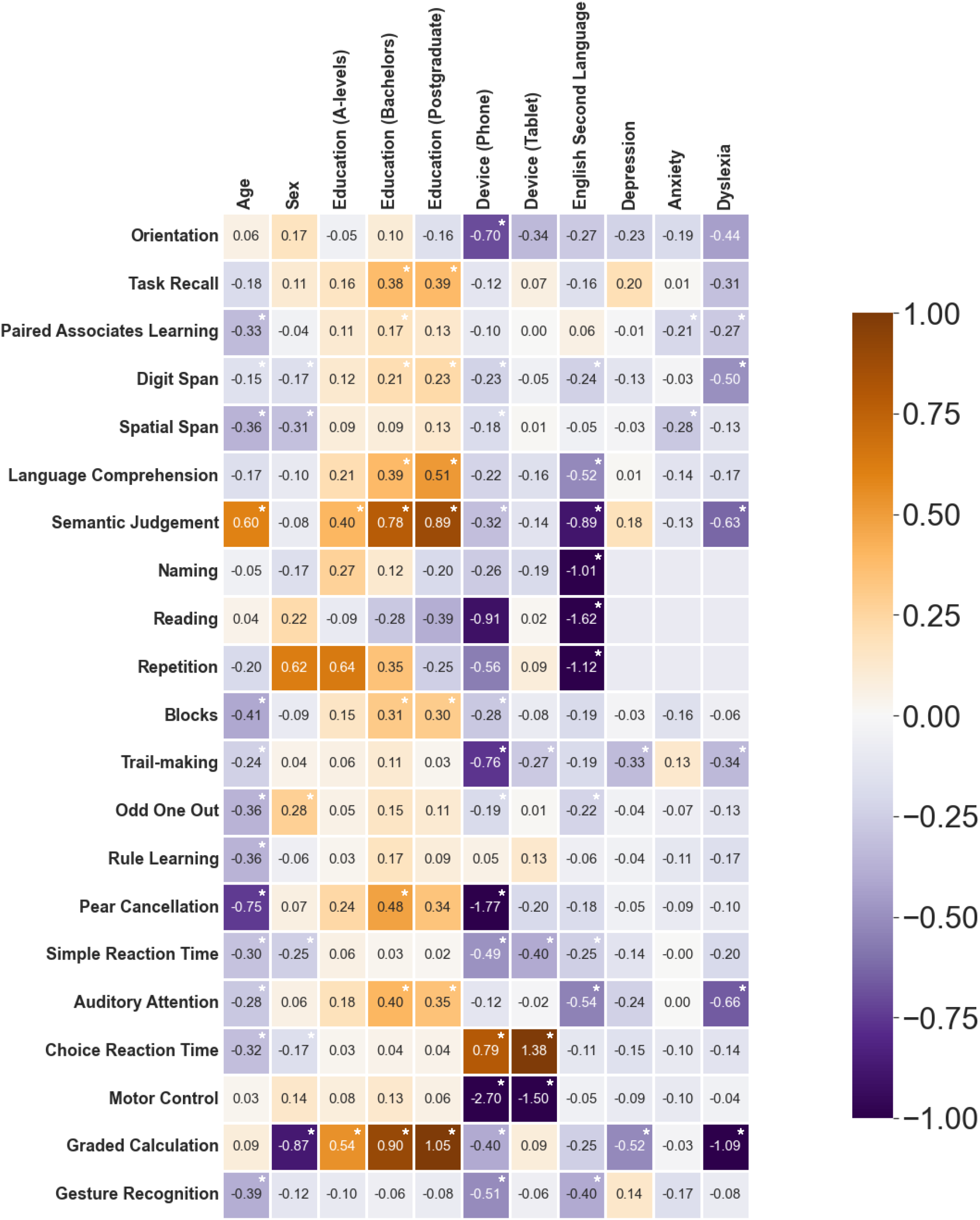
Relationship between cognitive performance and 8 confounding factors in the large normative sample, quantified via standardised regression coefficients. The regression reference categories for Education and Device were ‘≤GCSE-educated’ and ‘Desktop Computer’, respectively. Negative coefficients (cool colours) indicate the covariate was associated with worse cognitive performance. Empty cells represent coefficients not used in the modelling for the optional speech production tasks given the smaller normative sample. Regression coefficients for tasks with normally distributed data are in standard deviation units, and those with non-gaussian distribution are in log-odd units. Positive coefficients are indicative of better performance. ‘*’ = Uncorrected statistically significant predictors.

### Reliability of IC3 technology

#### Internal consistency amongst tasks

The task-level internal consistency was generally good, particularly for tasks with high variability on the primary outcome measure, with 17/21 tasks showing split-half alpha values surpassing the standard threshold for good internal consistency (α=0.70). See Supplementary Material 5.3. for more detailed methodology. Nevertheless, a small subset of tasks exhibited lower alpha values (i.e., Orientation, Task Recall, Gesture Recognition, and Calculation, marked with ‘†’ in Supplementary Table 2). A well-known limitation of the split-half reliability is its dependency on a large number of trials, with shorter tasks having inherently low reliability estimates.^20^ Thus, the low alpha values may be attributed to the low number of trials (4-6) in these short screening tasks, despite the tasks being preferable in clinical settings due to their lower burden on patients. A further factor is the ceiling effect in these short tasks such that, a single error results in a significant change in the trial’s relative ranking and estimated covariance between trials.

#### High test-retest reliability across time

There was no significant difference (FDR corrected) in performance between two sessions on any of the IC3 tasks with strong equivalence across all measures in sub-cohort A (Supplementary Material 5.4.). There was moderate to high correlation between sessions for tasks with high variance within the group, and smaller correlation for those with low variance due to ceiling effects despite similar group means across sessions. When diurnal associations with performance were examined across inter-session time interval and that of time of day when the assessment was performed, the results showed that these time-related factors did not explain change in cognitive scores across the two sessions (Supplementary Material 5.5.). Overall, these results demonstrated a stable performance of the control group on IC3 across time.

#### The remote monitoring technology shows equivalent performance between unsupervised and supervised testing environment

With the exception of two motor tasks (Choice Reaction Time and Motor Control), there were no differences between performance of supervised and unsupervised testing environment. (Supplementary Material 5.6.). Given that the performance on the two motor tasks was strongly dependent on the device type used (Figure 2), additional Bayesian regression modelling of these two tasks were conducted with device and environment as predictors, which confirmed that the session associations for these tasks were driven not by the supervised/unsupervised environment but by the device used to perform the assessment. (Supplementary Material 5.6.). Thus, we conclude that there were no significant direct effects of conducting assessment within an unsupervised testing environment.

#### The online technology is robust to learning effects across four timepoints

We examined the potential effect of learning by analysing performance across repeated testing timepoints in 30 controls (sub-cohort C) who completed the IC3 four times over the course of two weeks. The results showed that IC3 assessment has minimal learning effects within this expected testing time interval (detailed analyses shown in Supplementary Material 5.7.).

### IC3 technology in patients with stroke

#### The online battery is sensitive to group differences across all tasks

Data from 90 patients with stroke were analysed. The IC3 testing was performed during the acute post-stroke phase in 68% (3±3 days [median±IQR] post stroke) and in the sub-acute/chronic post-stroke phase in 32% (115±210 days post stroke). See Supplementary Materials 5.1. and 5.2.

The task level average group performance was calculated in ‘deviation from expected’ standard deviation units as described above. Healthy controls significantly outperformed patients across all tasks, showing moderate-to-large effect sizes in the majority of tasks after accounting for eight confounding factors (19/21 tasks, *P*<0.05 FDR corrected; d>0.43 for all tasks). See Figure 4 and Supplementary Material 5.8. for detailed statistical results.

**Figure 4:**
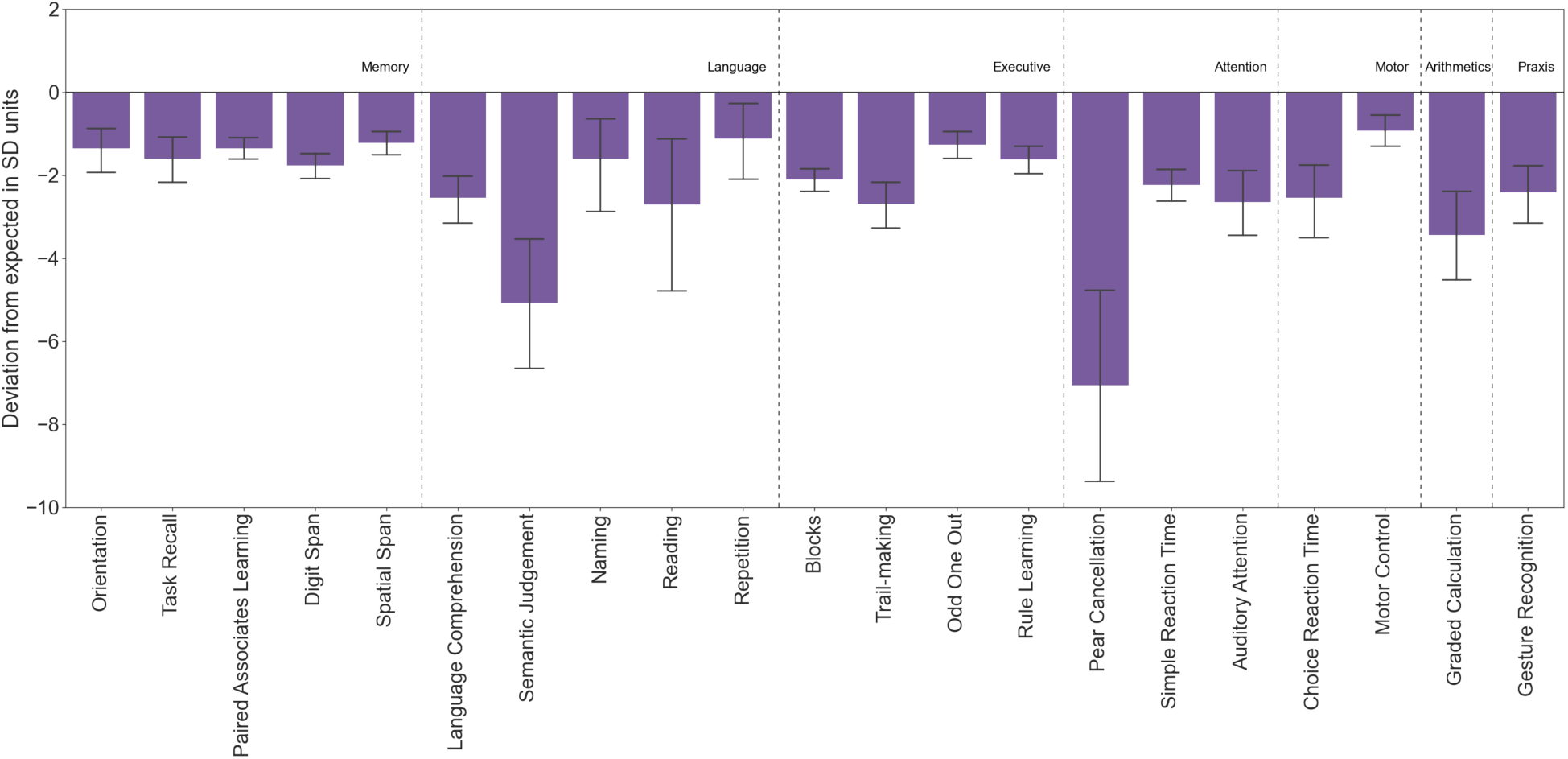
Patients with stroke had significantly worse performance than controls (FDR corrected). Error bar= 95% CI. 0= mean control performance.

#### The online battery correlates with clinical neuropsychological scores and functional impairment after stroke

A data driven ‘IC3 global composite score’, was derived from factor analysis based on combined data from patients and controls (Supplementary Material 5.9.). This had a positive loading on individual tasks and accounted for 47% of the variance (Figure 5A). Six group factors, each loading on a subset of tests were also derived, intuitively mapping onto Executive Function (F1), Language/Numeracy (F2), Working Memory (F3), Attention (F4), Motor Ability (F5) and Memory Recall (F6). The factor analysis fit was robust (CFI=0.94, RMSEA=0.01) with good internal consistency (omega=0.79).

**Figure 5.**
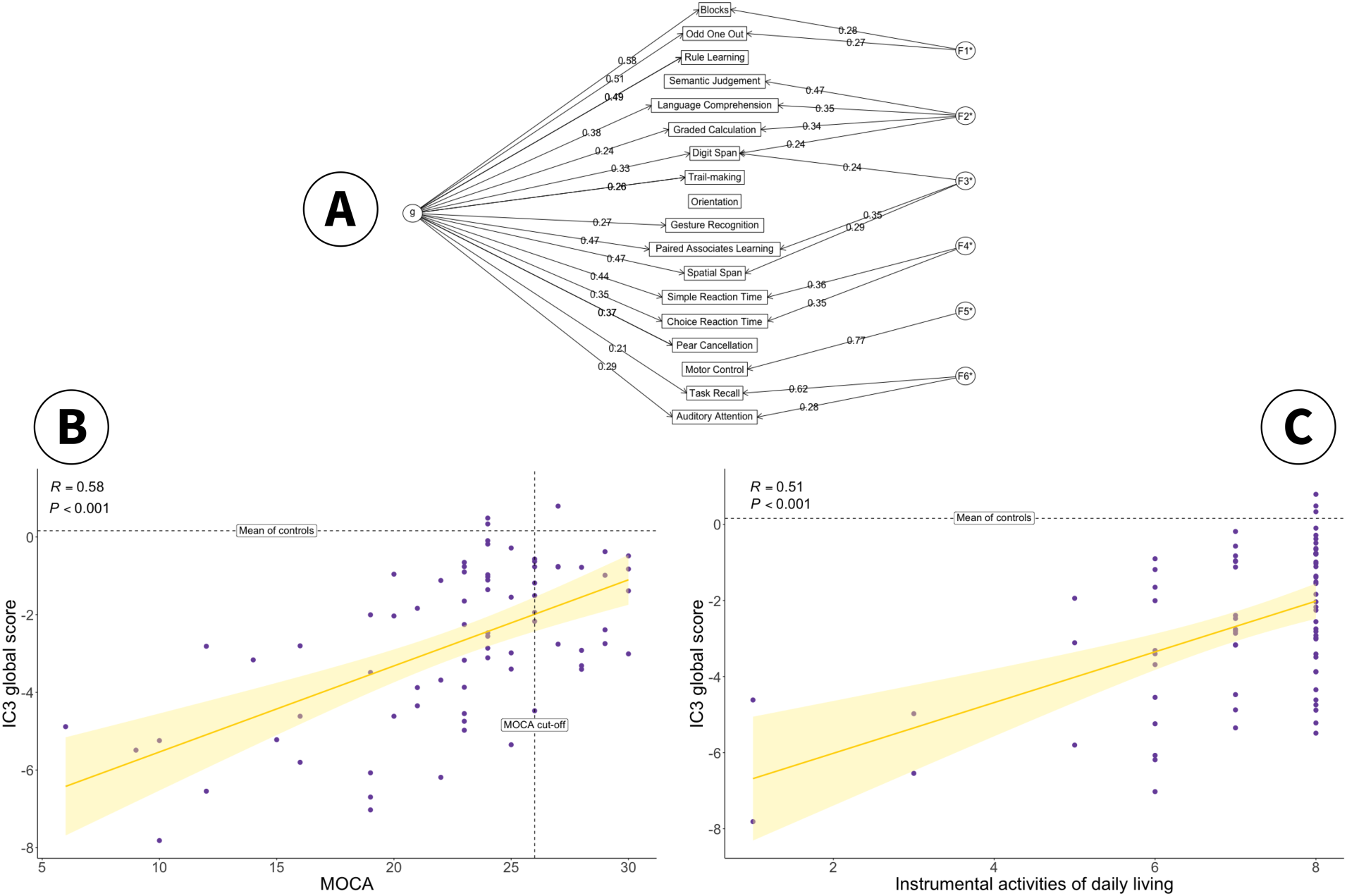
**A.** The solution to a bifactor exploratory factor analysis on the combined dataset of patients and controls. The global cognitive measure for the IC3 is defined as the ‘g’ factor. The remaining 6 factors intuitively map to Executive Function (F1), Language/Numeracy (F2), Working Memory (F3), Attention (F4), Motor Ability (F5) and Memory Recall (F6). Figure 5B. Correlation between IC3 global score (g) and clinically validated total MoCA score in patients (N=80). Vertical dotted line: MoCA cut-off for normality. Horizontal dotted line: mean global IC3 in controls. Shaded area represents 95% Confidence Interval. Figure 5C. Relationship between IC3 global score (g) and post-stroke quality of life metrics (IADL). Horizontal dotted line: mean global IC3 in controls. Error bar represents 95% Confidence Interval.

In keeping with task-level results, IC3 global score (g) was significantly lower in patients compared to controls (P<.001, Supplementary Material 5.9.). Furthermore, the IC3 global score and total MoCA scores were significantly correlated in patients (r(78)=0.58, R^2^=0.33, *P*< 0.001, Fig. 5B), indicating that the IC3 performance maps onto a clinically validated neuropsychological screen.

To assess the external validity of IC3, IC3 global performance was also related to functional impairment after stroke as defined by the Instrumental Activities of Daily Living (IADL) score.^15^ Worse global cognitive performance on the IC3 was associated with worse functional impairment after stroke (r(78)=0.51, R^2^=0.26, *P*<0.001, Fig. 5C).

Conversely, MoCA had a considerably weaker relationship with functional deficits post-stroke, explaining approximately half of the variation explained by the IC3 (r(78)=0.38, R^2^=0.14, *P*< 0.001). A separate linear regression analysis was conducted to quantify this difference, with MoCA and IC3 as independent predictors, and IADL as the dependent variable. The results show that there was no longer a main effect of MoCA on functional impairment when accounting for the IC3 global score (*P*=.22), whilst the IC3 global score remained highly significant (*P*=.004). Moreover, the inclusion of MoCA only explained an additional 1% of variance, suggesting that MoCA does not bring any additional information beyond what is captured by the IC3.

Furthermore, we demonstrate strong divergent validity for IC3 (Supplementary Material 5.10.), as shown by an expected lack of correlation between cognitive performance in patients and variables known not to be related to cognition (i.e. admission cholesterol levels).

#### The online monitoring technology is more sensitive to mild cognitive impairment than standard clinical assessment

IC3 showed high sensitivity at both the domain level and the task-level (as shown in high true positives in dark purple, Figure 6A). Given that IC3 was specifically designed to detect mild impairment, the sensitivity of the MoCA screening tool was also assessed against the IC3 and found to be weaker (Figure 6A, in yellow). A chi-square test indicated that this difference is statistically significant at p<0.001. See Supplementary Material 5.11. for details on how the ‘ground truth’ and domains were chosen.

**Figure 6.**
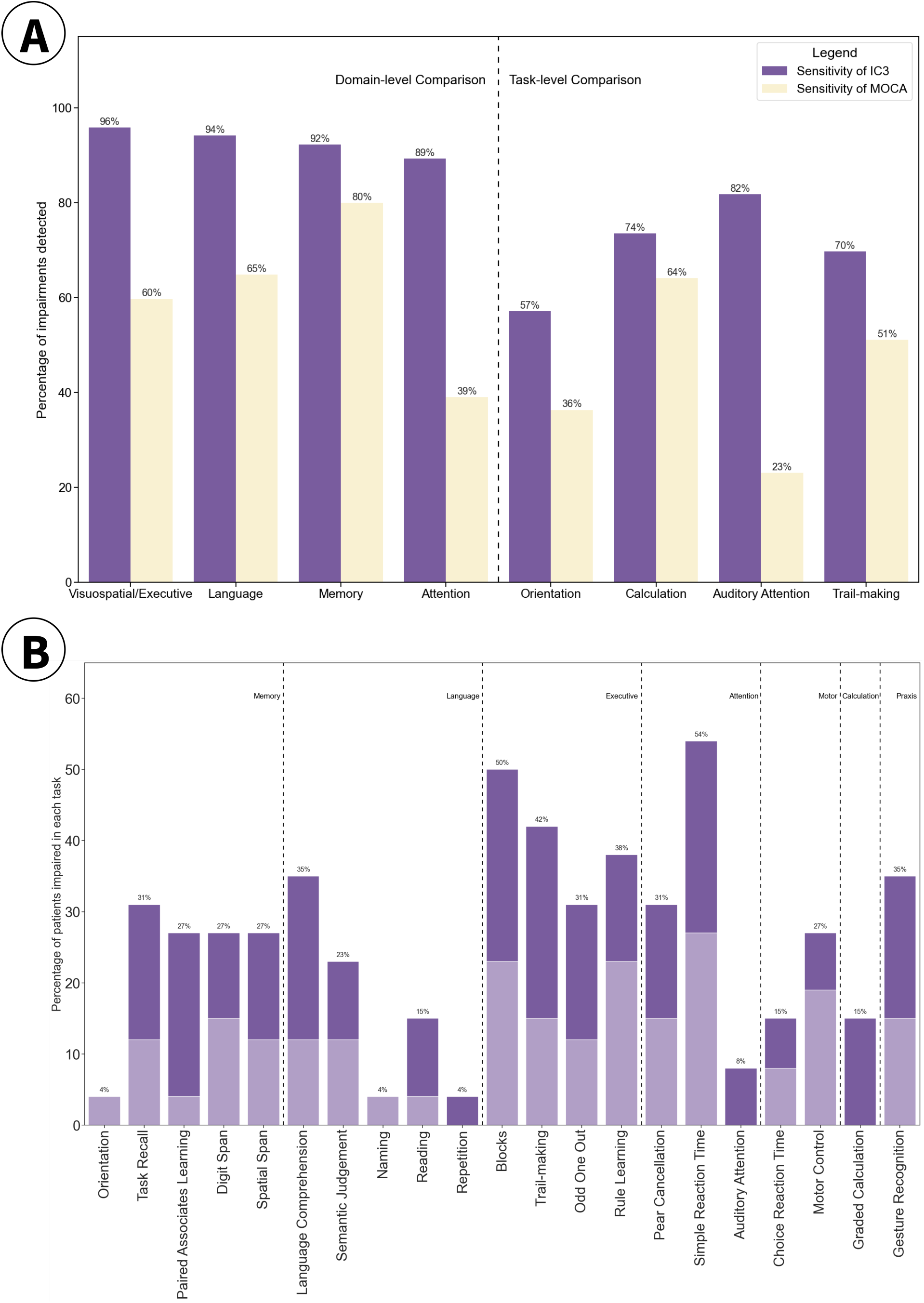
**A** Sensitivity of IC3 against MoCA (purple) and vice versa (yellow) in detecting impairment at domain and task level where a clear 1:1 mapping was present. Figure 6B. Percentage of patients classed as impaired on the IC3, within a sub-group of stroke patients that were deemed cognitively healthy on the MoCA (N=27). Dark purple= scores from the acute stage of the stroke. Lighter purple= sub-acute/chronic stage.

In addition, as shown in Figure 6B, IC3 was able to detect a substantial proportion of impairment in patients, even when those patients were classed as ‘healthy’ according to their MoCA performance (score ≥26/30). These impairments were detected in both the acute (dark purple) and the sub-acute/chronic stage (light purple) after stroke, highlighting IC3’s ability to detect mild impairment, undetected by clinical screens, in all stages of recovery after stroke.

## Discussion

The current study evaluated the IC3 battery, a novel digital online cognitive testing technology designed to enable large-scale identification and monitoring of cognitive sequalae after stroke and related vascular disorders. The robustness and reliability of the battery was extensively demonstrated and reinforced through high test-retest reliability, equivalent performance between supervised and non-supervised settings, minimal learning effects across multiple timepoints and high psychometric validity. An extensive normative sample of more than 6,000 older adults from the United Kingdom, age-matched to the stroke population, was systematically gathered and leveraged to calculate patient-specific prediction and impairment scores. While accounting for a wide range of demographic and neuropsychiatric variables, IC3 was able to differentiate healthy controls and stroke survivors across all tasks on the IC3 battery, with effect sizes ranging from moderate to very large (d=0.43-1.59).

Importantly, patient outcomes derived from IC3 demonstrated strong concordance with results from clinical scores available in the patients (MoCA), but with superior sensitivity when detecting mild cognitive impairments and stronger correlation with patient-reported functional impairments post-stroke (IADL). Reassuringly, the convergence validity of the IC3-derived outcomes in patients was balanced against an expected good divergent validity as shown with no discernible correlation with admission cholesterol levels. Collectively, these findings provide compelling evidence supporting the validity of remote digital testing via the IC3 platform as a clinical tool for assessing and monitoring cognition following stroke and related vascular disorders.

Compared to other assessment tools, IC3 battery provides: 1-scalability and cost-effectiveness in post-stroke cognitive monitoring in both healthcare and research settings, 2-sensitivity to mild cognitive impairments, 3-specificity to deficits found post-stroke, meeting the national guidelines requirements for cognitive assessment in stroke, 4-nuanced response metrics per individual tests (e.g., accuracy, reaction time and trial-by-trial variability), and 5-the ability to output automated real-time patient predictive scores accounting for confounding demographic factors.^5,7,8,21^

Currently available stand-alone cognitive screening tools that are commonly used in routine clinical care mostly are either not stroke-specific, or not comprehensive. Prominent examples include the MoCA, Mini Mental State Examination, and Addenbrooke’s Cognitive Examination-Revised, which are tailored to detect deficits in neurodegenerative dementias and are not tailored to patients with stroke, who often have domain-specific as well as domain-general deficits.^9,22,23^ Additionally, increasingly used cognitive screens designed specifically for stroke, such as the OCS, although more sensitive than MoCA, are not comprehensive enough to allow a deep cognitive phenotyping and miss the milder end of the severity spectrum.^24^ The digital OCS-Plus only assesses memory and executive function and requires a trained staff to administer, thus limiting its scalability and affordability compared to the IC3.^18^ Other digital platforms such as CANTAB, usually used in the setting of research into neurodegenerative/ psychiatric disorders, are not stroke-specific nor able to provide patient-predictive scores.^25^ The IC3 assessment battery developed and applied in this study addresses these shortcomings, building the foundation for routine detailed monitoring of cognition in healthcare setting, and for scalable large-scale population-based studies of post-stroke cognitive impairment.

The evidence presented in this paper is further corroborated by recent studies from our groups, showing the feasibility of online cognitive testing for identification and monitoring of impairments in neurological disorders, such as traumatic brain injury and autoimmune limbic encephalitis.^17,26^ Collectively, these studies have shown that online assessments correlate well with standard clinical evaluations while exhibiting higher sensitivity, essentially providing a hyper-resolution on cognitive deficits relative to common scales. The current results not only align with these findings, but also provide much more extensive reliability and validity metrics, showcasing the 1) robustness of the IC3 assessment, 2) its superiority against standard clinical screens and 3) its ability to predict functional outcomes after stroke. These results provide supporting evidence for integration of such platforms in clinical care pathway, providing additional insight into clinical decisions; for instance, on when to perform, potentially costly, imaging or in-person testing during the follow-up period of the disease, and inform rehabilitation decisions.

Nevertheless, it is important to consider the current findings in the context of certain limitations. Similar to most standard pen-and-paper tests, the IC3 tasks were administered in a predetermined order. Consequently, missing data were more likely to occur for tasks towards the latter part of the battery, potentially leading to underrepresenting participants with higher level of impairments for these tasks. However, it is worth noting that this phenomenon is inherent in all cognitive assessment methodologies, and the inability to complete the assessment can itself be considered a meaningful outcome measure. In the current study, only patients who had completed at least 50% of the IC3 assessment were included in the analysis, and the last task of the IC3 normative cohort had 4782 respondents surpassing the normative sample size for most conventional assessment tools.

We showed that the IC3 has superior sensitivity to MoCA. Here MoCA was used as the approximate ground truth for detecting post-stroke impairments, as this was routinely available as a clinical screening test in our patients. However, MoCA is neither stroke-specific nor sensitive to impairment when compared to in-depth neuropsychological evaluation.^27,28^ In the absence of such data in our patient cohort, and the likelihood that MoCA missed milder impairments, it was deemed inappropriate to assess the specificity or false positive rate of IC3 against MoCA. This problem has been previously noted in digital cognitive tests that evaluate their efficacy against MoCA.^29^ Future work, assessing the sensitivity of MoCA against more sensitive stroke-specific tests are in progress.

To maximise engagement of more severely impaired patients, the IC3 allows for unlimited number of breaks to be taken at the end of each task and provides built-in task/trial skipping functions to minimise fatigue in these patients. Nevertheless, these patients will not be suitable for unsupervised testing given their well-documented issues with engagement.^30^ The unsupervised administration is most likely appropriate for patients with mild-to-moderate impairment, who have the highest potential for regaining independence and who may benefit most from personalised treatments and rehabilitation.

Due to its cost-effectiveness and scalability, IC3 can be further developed for wide adoption as a clinical diagnostic and monitoring tool for patients with stroke and related vascular disorders. This will be tested in clinical trials within the healthcare setting. Such implementation will facilitate the detection and longitudinal monitoring of cognitive impairment after stroke at minimal cost. Given its sensitivity to mild impairment, IC3 can be used as the main cognitive outcome measure in clinical research studies in patients with stroke. We are currently adopting this approach in a longitudinal observational study of post-stroke cognition alongside blood biomarkers and brain imaging to identify mechanisms of recovery following stroke.^10^ It is anticipated that these findings will inform tailored, personalised rehabilitation strategies for more effective recovery, a prospect not achievable with current assessment methodologies.

## Acknowledgements

We are grateful to all the patients and participants for their time and involvement in the study. We thank Sabia Combrie and Charlotte Barot for their contribution patient data collection, Joseph Coghlin for his help with control data collection, Ziyuan Cai and Georgia Gvero for manually transcribing the speech data. We are grateful to Daniel Alexandre Romano Alves for advising the design of the IC3 graphics.

## Data availability

A subset of the datasets used can be made available on reasonable request from the corresponding author and upon institutional regulatory approval.

## Code availability

The code used in the pre-processing and group-level statistical analysis for this study will be made available on publication. The underlying code for the task designs is not publicly available for proprietary reasons.

## References

1. Douiri A, Rudd AG, Wolfe CD. Prevalence of poststroke cognitive impairment: South London stroke register 1995–2010. Stroke. 2013;44(1):138–45.

2. Hamilton OKL, Backhouse EV, Janssen E, Jochems ACC, Maher C, Ritakari TE, et al. Cognitive impairment in sporadic cerebral small vessel disease: A systematic review and meta-analysis. Alzheimer’s & Dementia. 2021;17(4):665–85.

3. Stolwyk RJ, Mihaljcic T, Wong DK, Hernandez DR, Wolff B, Rogers JM. Post-stroke Cognition is Associated with Stroke Survivor Quality of Life and Caregiver Outcomes: A Systematic Review and Meta-analysis. Neuropsychol Rev. 2024 Mar 11

4. Stolwyk RJ, Mihaljcic T, Wong DK, Chapman JE, Rogers JM. Poststroke Cognitive Impairment Negatively Impacts Activity and Participation Outcomes. Stroke. 2021 Feb;52(2):748–60.

5. McMahon D, Micallef C, Quinn TJ. Review of clinical practice guidelines relating to cognitive assessment in stroke. Disability and Rehabilitation. 2022 Nov 20;44(24):7632–40.

6. Hill G, Regan S, Francis R, Mead G, Thomas S, Salman RAS, et al. Research priorities to improve stroke outcomes. The Lancet Neurology. 2022 Apr 1;21(4):312–3.

7. Party ISW. National clinical guideline for Stroke for the United Kingdom and Ireland. London: Royal College of Physicians. 2023;

8. Quinn TJ, Richard E, Teuschl Y, Gattringer T, Hafdi M, O’Brien JT, et al. European Stroke Organisation and European Academy of Neurology joint guidelines on post-stroke cognitive impairment. European Stroke Journal. 2021 Sep;6(3):I–XXXVIII.

9. Nasreddine ZS, Phillips NA, Bédirian V, Charbonneau S, Whitehead V, Collin I, et al. The Montreal Cognitive Assessment, MoCA: A Brief Screening Tool For Mild Cognitive Impairment. Journal of the American Geriatrics Society. 2005;53(4):695–9.

10. Gruia DC, Trender W, Hellyer P, Banerjee S, Kwan J, Zetterberg H, et al. IC3 protocol: a longitudinal observational study of cognition after stroke using novel digital health technology. BMJ Open. 2023 Nov 1;13(11):e076653.

11. Hampshire A. Great british intelligence test protocol. 2020;

12. Marin RS, Biedrzycki RC, Firinciogullari S. Reliability and validity of the apathy evaluation scale. Psychiatry Research. 1991 Aug 1;38(2):143–62.

13. Krupp LB, LaRocca NG, Muir-Nash J, Steinberg AD. The fatigue severity scale: application to patients with multiple sclerosis and systemic lupus erythematosus. Archives of neurology. 1989;46(10):1121–3.

14. Herrmann N, Mittmann N, Silver IL, Shulman KI, Busto UA, Shear NH, et al. A validation study of The Geriatric Depression Scale short form. International Journal of Geriatric Psychiatry. 1996;11(5):457–60.

15. Lawton MP, Brody EM. Assessment of older people: self-maintaining and instrumental activities of daily living. The gerontologist. 1969;9(3_Part_1):179–86.

16. Hampshire A, Azor A, Atchison C, Trender W, Hellyer PJ, Giunchiglia V, et al. Cognition and Memory after Covid-19 in a Large Community Sample. New England Journal of Medicine. 2024 Feb 29;390(9):806–18.

17. Del Giovane M, Trender WR, Bălăeţ M, Mallas EJ, Jolly AE, Bourke NJ, et al. Computerised cognitive assessment in patients with traumatic brain injury: an observational study of feasibility and sensitivity relative to established clinical scales. EClinicalMedicine. 2023;59.

18. Demeyere N, Haupt M, Webb SS, Strobel L, Milosevich ET, Moore MJ, et al. Introducing the tablet-based Oxford Cognitive Screen-Plus (OCS-Plus) as an assessment tool for subtle cognitive impairments. Sci Rep. 2021 Apr 12;11(1):8000.

19. Hartshorne JK, Germine LT. When Does Cognitive Functioning Peak? The Asynchronous Rise and Fall of Different Cognitive Abilities Across the Life Span. Psychol Sci. 2015 Apr 1;26(4):433–43.

20. Pronk T, Molenaar D, Wiers RW, Murre J. Methods to split cognitive task data for estimating split-half reliability: A comprehensive review and systematic assessment. Psychon Bull Rev. 2022 Feb 1;29(1):44–54.

21. Giunchiglia V, Gruia D, Lerede A, Trender W, Hellyer P, Hampshire A. Iterative decomposition of visuomotor, device and cognitive variance in large scale online cognitive test data. Preprint. 2023.

22. Kurlowicz L, Wallace M. The mini-mental state examination (MMSE). Vol. 25, Journal of gerontological nursing. 1999. p. 8–9.

23. Mioshi E, Dawson K, Mitchell J, Arnold R, Hodges JR. The Addenbrooke’s Cognitive Examination Revised (ACE-R): a brief cognitive test battery for dementia screening. International Journal of Geriatric Psychiatry. 2006;21(11):1078–85.

24. Demeyere N, Riddoch MJ, Slavkova ED, Bickerton WL, Humphreys GW. The Oxford Cognitive Screen (OCS): Validation of a stroke-specific short cognitive screening tool. Psychological Assessment. 2015 Sep;27(3):883–94.

25. Smith PJ, Need AC, Cirulli ET, Chiba-Falek O, Attix DK. A comparison of the Cambridge Automated Neuropsychological Test Battery (CANTAB) with “traditional” neuropsychological testing instruments. Journal of Clinical and Experimental Neuropsychology. 2013 Mar 1;35(3):319–28.

26. Shibata K, Attaallah B, Tai XY, Trender W, Hellyer PJ, Hampshire A, et al. Remote digital cognitive assessment reveals cognitive deficits related to hippocampal atrophy in autoimmune limbic encephalitis: a cross-sectional validation study. eClinicalMedicine. 2024 Feb 2

27. Chan E, Khan S, Oliver R, Gill SK, Werring DJ, Cipolotti L. Underestimation of cognitive impairments by the Montreal Cognitive Assessment (MoCA) in an acute stroke unit population. Journal of the Neurological Sciences. 2014 Aug 15;343(1):176–9.

28. Demeyere N, Riddoch MJ, Slavkova ED, Jones K, Reckless I, Mathieson P, et al. Domain-specific versus generalized cognitive screening in acute stroke. J Neurol. 2016 Feb 1;263(2):306–15.

29. Roberts R, Vohora R, Webb SS, Demeyere N. Validating the OCS-Plus against a clinical standard: A brief report. Journal of Neuropsychology. 2024 Apr 11

30. Bill O, Zufferey P, Faouzi M, Michel P. Severe stroke: patient profile and predictors of favorable outcome. Journal of Thrombosis and Haemostasis. 2013 Jan 1;11(1):92–9.

