## Supplementary Material for "Online monitoring technology for deep phenotyping of cognitive impairment after stroke"

Supplementary Materials

### 1. **IC3 Task Descriptions**

IC3 is composed of 22 tasks (18 short cognitive tasks with additional 4 optional speech production tasks) with additional demographic, health and psychiatric questionnaires which are described below. IC3 can be completed under 1 hour and has built-in automated break reminders every 20 minutes, with the additional option to take an unscheduled break at any time.

The tasks are grouped based on the primary cognitive domain intended to be probed. All task instructions are presented both visually and audibly to aid understanding in patients with aphasia. Examples of task stimuli presentation are shown in Figure 1 and the mean, standard deviation (STD) median, inter-quartile range (IQR), of the normative cohort is presented alongside the primary outcome in each task.

#### 1.1. **Memory domain**

##### 1.1.1. Orientation

Orientation provides a measure of long-term memory, and orientation to time is a strong predictor of subsequent cognitive decline in the elderly.^1^ Here participants are required to answer 4 multiple-choice questions in response to orientation to time (year, month, week, and time of day). Each question is, by default, presented both visually and audibly, and there is also the option for auditory presentation of forced-choice options. Participants must make responses within 20 seconds per question.

**Primary outcome.** Number of correct responses out of 4 (N=6279, Mean=3.93, STD=0.27, Median=4.0, IQR=0.0, Range=2.0-4.0).

**Secondary outcome.** Mean reaction time (RT) during correct responses (milliseconds).

##### 1.1.2. Task Recall

Task Recall provides a measure of short-term memory via cued recall. At the end of the cognitive battery, participants are shown stimuli from previously completed tasks, along with foils, and asked to identify those that have been presented before. The task has four trials each with 15 seconds maximum response time per trial. Each question is, by default, presented both visually and audibly. This task contains two different versions, which are alternated on repeated administration of the cognitive battery to minimise learning effects. The two versions are matched for task instructions, number of trials, and tasks probed, but have different target and foil stimuli.

**Primary outcome.** Number of correct responses out of 4 (N=4782, Mean=3.78, STD=0.45, Median=4.0, IQR=0.0, Range=2.0-4.0).

**Secondary outcome.** Mean RT during correct responses.

##### 1.1.3. Paired Associates Learning (PAL)

PAL provides a measure of associative working memory required for learning. This type of learning has been shown to be impaired early on in neurodegenerative dementias such as Alzheimer's Disease.^2^ In this task, abstract symbols are presented in sequence for 1 second each, randomly positioned within a 4 by 4 grid. Participants are instructed to remember the symbol-location associations. Following the sequence presentation, participants are asked to indicate, one at a time, where each symbol was located on the grid. If a mistake is made, the same sequence of symbols is presented again, and they are given up to 3 times to remember it. The number of items in the sequence increases one at a time after each correct sequence completion. The tasks stops either after (a) the participant makes errors on more than two unique sequences, (b) a maximum of 9 symbol-location pairs are completed, or (c) 3 minutes have passed.

**Primary outcome.** Maximum number of symbol-location associations remembered (N=5721, Mean=5.38, STD=1.38, Median=5.0, IQR=2.0, Range=2.0-9.0).

**Secondary outcome**. Total number of errors made across all symbol-location associations.

##### 1.1.4. Digits Span

Digits Span provides a measure of verbal working memory. In this task, a set of numbers are presented in sequence, for 1 second each. Participants are instructed to remember the numbers in the order of presentation (forward span), and to indicate their responses on a keypad. In the first trial of the task, the participant is only required to recall a sequence of two digits, with the length of the sequence increasing using a ratchet system, where every time a sequence is recalled correctly, the length of the subsequent sequence is incremented by one. The digits are randomly generated, with a rule that there cannot be more than 2 consecutive digits presented in order one after the other. The participants are allowed only 2 attempts per sequence length. The task ends after the participant makes errors on two consecutive trials or after 3 minutes of play time.

**Primary outcome.** Maximum digit sequence length recalled (N=5279, Mean=7.24, STD=1.35, Median=7.0, IQR=2.0, Range=3.0-12.0).

**Secondary outcome**. Total number of errors made.

##### 1.1.5. Spatial Span

Spatial Span is the non-verbal analogue of the digit span and provides a measure of spatial working memory. Participants are shown a 4x4 grid. One at a time, in sequence, randomly chosen grid cells flash grey. Participants are instructed to recall the correct order in which the grid cells flash grey. The sequence length increases with each correct trial and the task progression criteria is identical to that of digit span task.

**Primary outcome.** Maximum digit sequence length recalled (N=5867, Mean=5.6, STD=1.13, Median=6.0, IQR=1.0, Range=3.0-10.0).

**Secondary outcome**. Total number of errors made.

#### 1.2. **Language Domain**

##### 1.2.1. Naming

This confrontation picture naming task provides a measure of object recognition and single-word naming ability. Participants are shown 30-line drawings of objects presented on a white background, and are asked to name them overtly. The list of stimuli is from the 30-item version of the Boston Naming Task.^3^ Each naming trial lasts a maximum of 15 seconds. Participants are given the option to skip the task if they do not wish their voice to be recorded and can also skip a naming trial by pressing a forward button. The trial order is randomised to minimise order effects.

**Primary outcome.** Proportion of pictures correctly named (semantically) out of 30. Dysarthric and dyspraxic errors are not penalised (N=130, Mean=0.96, STD=0.06, Median=0.97, IQR=0.07, Range=0.63-1.0).

**Secondary outcomes**. Global average score incorporating semantic, phonology, dysfluency, and self-correction errors, where each measure is scored out of 2 to give a maximum average score of 2 per naming item.

##### 1.2.2. Picture Description

The picture description task provides a measure of spontaneous speech production and fluency. Participants are shown 2 scenes (line drawings) and are asked to describe them in as much detail as they can using full grammatical sentences. The participants have up to 90 seconds to describe each picture but have the option to skip the task at any time.

**Primary outcome.** Total correct words per minute (Tokens) for both pictures. Normative ranges will be discussed in a separate paper due to the labour-intensive nature of scoring speech production.

**Secondary outcomes**. Average words per minute across the two pictures, semantic errors, phonological errors, dysfluencies.

##### 1.2.3. Reading

The reading task provides a measure of single-word lexical and non-lexical processing. Participants are shown 11 words, including 3 non-words and 4 function words, and are asked to name them overtly. Each trial lasts a maximum of 15 seconds. Participants are given the option to skip the task if they do not wish their voice to be recorded. The trial order is randomised to minimise order effects.

**Primary outcome.** Proportion of words correctly read (phonologically) out of 11 (N=130, Mean=0.99, STD=0.03, Median=1.0, IQR=0.0, Range=0.82-1.0).

**Secondary outcomes**. Global average score incorporating phonology, dysfluency, and self-correction errors, where each measure is scored out of 2 to give a maximum average score of 2 per reading item marked.

##### 1.2.4. Repetition

The repetition task provides a measure of single-word repetition ability. Participants hear a sequence of 20 words and are asked to repeat each individual word overtly. Each trial lasts a maximum of 15 seconds. Participants are given the option to skip the task if they do not wish their voice to be recorded. The words are selected from a condensed pool of words in the repetition task of the PALPA 9 that has been reduced further to 40 using a data driven approach.^4,5^ This reduced list is further divided into 4 categories based on phoneme count (low vs high) and their relative frequency (low vs high). The 20 words are randomly selected from each of the four categories (5 in each category) to minimise learning effects on repeat trials.

**Primary outcome.** Proportion of words correctly repeated (phonologically) out of 20, (N=130, Mean=0.99, STD=0.02, Median=1.0, IQR=0.0, Range=0.88-1.0).

**Secondary outcomes**. Global average score incorporating phonology, dysfluency, and self-correction errors, where each measure is scored out of 2 to give a maximum average score of 2 per reading item marked.

##### 1.2.5. Language Comprehension

The language comprehension task provides a measure of graded word and sentence comprehension. A target word or phrase is presented at the top of the screen, with a choice of four pictorial representations below. Participants must select the picture that best represents the target word or phrase. Each target word or sentence is presented both visually and audibly by default to aid comprehension. The task is designed to increase in difficulty after four correct responses or once all stimuli in that task level are presented. There are three difficulty levels in the task: 1) maximum of 8 single-word picture-matching where an inanimate target word has to be matched to the correct picture amongst 3 foils (phonological distractor, semantic distractor and unrelated distractor); 2) maximum of 6 simple phrase picture-matching where short relational phrases (e.g. “bird in front of the nest”) are matched to the target picture amongst 3 foils that includes the reversed relational representation; and 3) maximum of 6 complex sentence picture-matching requiring verbal reasoning and complex syntax understanding (e.g., “the circle in the square is red”). The trial order within each task difficulty is randomised to minimise order effects. Each trial lasts a maximum of 15 seconds, and the task is designed to end prematurely if 6 consecutive errors are made.

**Primary outcome.** Number of correct responses (N=5025, Mean=18.67, STD=1.42, Median=19.0, IQR=2.0, Range=13.0-20.0).

**Secondary outcomes.** Total number of errors, number of semantic errors in level 1, number of phonological errors in level 1, and maximum level reached.

##### 1.2.6. Semantic Judgement

This task captures semantic understanding of written words. A target word (e.g., “forest”) is shown at the top of the screen, with a choice of 3 words below it: the target semantic match (“woods”), and two foils (“wine” and “boat”). The stimuli word list are derived from the synonym judgement task originally described by Jefferies and colleagues in 2009 and was further condensed to a list of 48 target words.^6,7^ There are 23 trials in total and each target word is presented visually and audibly to aid comprehension. The task has 6 graded-difficulty levels based on imageability and frequency of the target word, each containing up to 8 trials. An example of the first difficulty level containing trials of high frequency and high imageability words include “woods” and its semantic match “forest”. Low frequency and low imageability trials contain words such as “dirge” matched with “lament”. The participant moves up the difficulty category if they correctly respond to three consecutive trials and moves down the difficulty category with three consecutive incorrect answers. If they finish all the trials in the highest difficulty level, the task ends. The participant scores a maximum of 3 points for each correct trial within each of the first 5 difficulty categories. They score a maximum of 8 points for the final difficulty category. The trial order and the position of the target and foils on the screen are randomised to minimise order and learning effects respectively. Each trial lasts a maximum of 15 seconds, and the task is designed to end prematurely if 6 consecutive errors are made.

**Primary outcome.** Number of correct responses (N=4995, Mean=22.64, STD=0.78, Median=23.0, IQR=1.0, Range=16.0-23.0).

**Secondary outcomes.** Number of semantically related errors, and total number of errors.

#### 1.3. **Executive Function**

##### 1.3.1. Blocks

This task provides a measure of visuospatial reasoning. A 7*7 grid of coloured blocks is presented on the left. Participants must remove blocks from the grid such that the remaining blocks resemble a target grey silhouette grid on the right. The grids are colour-blind friendly for protanopia, the most common form of colour-blindness. A maximum of 15 trials can be completed, with increasing difficulty which is modulated by two factors; the number of blocks needed to be removed (between 1-3) and the number of unsupported blocks that must fall under gravity in order to reach the target silhouette (between 0-3). Each trial is randomly generated, thus minimising learning effects. Participants perform three practice trials to ensure task comprehension is achieved before progressing to the main task. Each trial lasts a maximum of 20 seconds, and the task is designed to end prematurely if 6 consecutive mistakes are made or if the participant fails all practice trials. The task lasts for a maximum of 2 minutes.

**Primary outcome.** Total number of correct trials (N=5039, Mean=10.03, STD=2.56, Median=10.0, IQR=4.0, Range=3.0-15.0).

**Secondary outcome.** Total number of errors across all trials, mean RT, mean RT during correct responses.

##### 1.3.2. Trail-making

This task provides a measure of processing speed and task switching. The task is split into three parts. During the first level, the participant is shown a collection of 7 stars and is instructed to press the stars in sequence according to the increasing number of features (spikes) displayed. In the second level, the same rules apply but participants are now shown 7 circles with increasing number of features (concentric elements within). During the third level, both circles and stars are displayed, and participants are instructed to follow the above-mentioned rules whilst alternating between the stars and the circles. The first and second levels contain practice trials to ensure adequate comprehension of task instructions. The screen location of the stars and circles is randomised, to minimise learning effects. If no responses are made for 15 seconds or the practice trials fail (>7 errors), the participant is reminded of the task instructions one more time, after which such errors will result in the task ending prematurely with a score of zero. The first and second level will also end if the participant cannot finish them within 1.5 minutes each. The third level ends after 2 minutes due to its increased complexity.

**Primary outcome.** Switch cost in errors, calculated as the difference in errors between the switching level (third level) and the combined non-switching levels, (N=5419, Mean=0.53, STD=1.13, Median=0.0, IQR=1.0, Range=0.0-6.0).

**Secondary outcome.** Switch cost in accuracy, switch cost in RT, number of errors in the switching level, RT in the switching level.

##### 1.3.3. Odd One Out

This task provides a measure of abstract reasoning, while also tapping into visuo-spatial processing. Participants are shown 9 panels that contain different abstract shapes. The panels differ based on colour, the type of shape, and the number of abstract items contained within them. Participants are instructed to identify the panel that is the ‘odd one out’. The trial difficulty is modulated through (1) the degree of similarity between the odd one out and the foils, (2) the number of unique foils and (3) whether the odd one out can be identified based on a single criteria (e.g., colour) or a combination of criteria (e.g. colour and shape). The stimuli for each trial are randomly generated, according to the above-mentioned difficulty criteria, thus minimising learning effects. Each trial lasts a maximum of 20 seconds, and participants are given 2 minutes to identify as many patterns as possible. The task may end early if participants make 6 consecutive errors. There participants complete a maximum of 18 trials.

**Primary outcome.** Number of correct responses (N=5672, Mean=11.6, STD=2.24, Median=12.0, IQR=3.0, Range=6.0-18.0).

**Secondary outcome.** Number of errors

##### 1.3.4. Rule finding

This task provides a measure of reinforcement learning, set shifting and task switching within and across dimensions. Participants are shown two abstract shapes and are instructed to identify the intended ‘correct’ one, through a process of trial and error where feedback is provided after each response. After correctly identifying the target shape for 6 consecutive trials, the ‘rule’ changes and the participants must switch to a new correct shape. There are a total of 9 different levels, with different shapes in each. The task is programmed to end after 2 minutes, regardless of how many levels have been achieved. The task will end early if more than 15 errors are made. This task contains two versions with identical instructions but different shapes, thus minimising learning effects on repeat administration of the task.

**Primary outcome.** Maximum level reached. Those who reach the final (9^th^ level) and finish all the trials in the allotted time are given a score of 10, (N=5485, Mean=8.02, STD=1.52, Median=8.0, IQR=1.0, Range=3.0-10.0).

**Secondary outcomes.** Switching errors. Mean duration across blocks.

#### 1.4. **Attention**

##### 1.4.1. Pear Cancellation

This task provides a measure of visual ability and spatial neglect. Participants are shown line drawings of pears and are asked to identify the complete pears amongst incomplete foils. Pears may be missing contours either on the left or on the right side. A practice block is completed at the start of the task to ensure task instruction comprehension. Failing the practice block automatically skips the remainder of the task. Participants have 3 minutes to identify all the complete pears.

**Primary outcome.** Proportion of correctly identified pears (N=6219, Mean=0.99, STD=0.03, Median=1.0, IQR=0.0, Range=0.8-1.0).

**Secondary outcomes.** Egocentric and allocentric neglect. Egocentric neglect is measured by whether participants miss targets (whole pears) on one side of the page. Allocentric neglect is measured by whether participants make false positive responses by cancelling an incomplete pear distractor where the contour is taken from one side.

##### 1.4.2. Simple Reaction Time

This task provides a measure of sustained visual attention. Participants are instructed to press a red target on the centre of the screen, as soon as it appears. The task lasts for maximum 3 minutes and the target is on the screen for 1 second, with an inter-stimulus gap of 0.5-2.5 seconds. The gap is randomly generated at the beginning of the task, to minimise learning effects.

**Primary outcome.** Mean RT during correct responses (N=5510, Mean=349.58, STD=59.46, Median=340.87, IQR=74.95, Range=211.85-680.07).

**Secondary outcomes. Total** Number of correct responses. Total number of timeouts (trials where the target appeared but no response was made). Total number of mis-clicks (trials where the participant made a response when there was no target on the screen).

##### 1.4.3. Auditory Attention

This task provides a measure of sustained auditory attention, working memory, target selection and inhibition. Participants are asked to press a red target in the centre of the screen every time they hear one of three high frequency words: ‘hi’, ‘come’, ‘up’. They are instructed to inhibit responses for their respective opposite foils (‘bye’, ‘go’, ‘down’). The 6 words are presented in two blocks each in random order, each being presented an equal number of times (3 presentations per word per block) with the inter-stimulus interval of a 2, 3 or 4 seconds gap during which participants can make a response for the preceding word. This period is randomly generated to minimise learning effects. The task lasts for a maximum of 2 minutes in total and will end earlier if there is no response for more than 10 consecutive trials.

**Primary outcome.** Number of correct responses (N=5757, Mean=35.19, STD=1.74, Median=36.0, IQR=1.0, Range=25.0-36.0).

**Secondary outcomes.** Mean RT during correct responses. Inhibition errors (responses on foils). Word-specific errors indicative of specific word encoding difficulties.

#### 1.5. **Motor ability**

##### 1.5.1. Choice Reaction Task

This task provides a measure of left-right hand motor dexterity and sustained attention. An arrow in the centre of the screen, points left or right. Participants are instructed to click on the side of the screen that the arrow points to as fast as possible. The arrow is presented on the screen for 1 second at a time with an inter-stimulus gap of 0.5-2.5 seconds; this period is randomly generated to minimise learning effects. The task has 60 trials and lasts for a maximum of 3 minutes.

**Primary outcome.** Number of correct responses (N=5181, Mean=56.96, STD=3.91, Median=58.0, IQR=4.0, Range=40.0-60.0).

**Secondary outcomes.** Number of incorrect trials. Number of timeouts (trials where the target appeared but no response was made). Number of mis-clicks (trials where the participant made a response when there was no target on the screen). Mean RT across correct trials.

##### 1.5.2. Motor Control

This task provides a measure of visuomotor co-ordination, multi-directional hand/arm motor dexterity and speed. A small moving red target appears on the screen and the participants are asked to click on it as soon as it appears. The target remains on the screen until it is selected. After it is selected the target re-appears in another randomly chosen location on the screen. The task lasts for a maximum of 3 minutes or until the participants completes 30 trials.

**Primary outcome.** Number of correctly identified targets defined as clicks within 25 pixels of the centre of the target (N=5208, Mean=27.9, STD=3.28, Median=29.0, IQR=3.0, Range=16.0-30.0).

**Secondary outcomes.** Number of incorrect responses. Mean RT during correct trials.

#### 1.6. **Numeracy**

##### 1.6.1. Calculations

This task provides a measure of simple arithmetic skills. Participants are shown a series of simple calculations involving addition or subtraction and are asked to select the correct answer from 4 multiple-choice options. The task increases in difficulty with each correct response, ranging from single digit to double digit addition/subtraction. Each trial lasts a maximum of 15 seconds. The task ends early if the participant makes 4 mistakes.

**Primary outcome.** Number of correct responses (N=5681, Mean=7.93, STD=0.4, Median=8.0, IQR=0.0, Range=4.0-8.0).

**Secondary outcomes.** Number of incorrect responses. Mean RT during correct responses.

#### 1.7. **Praxis**

##### 1.7.1. Gesture recognition

This task provides a digital proxy for ideomotor apraxia assessments. Previous studies have shown that performance on similar gesture recognition tasks are highly correlated with gesture production tasks.^8^ Participants are shown a series of 8 recorded gestures and are asked to identify the correct meaning from a choice of four written options, which can be further supported audibly if needed by pressing a loudspeaker sign. Half of the gestures are transitive, and the remainder are intransitive. Participants have a maximum of 25 seconds to answer each question during which time the video is played on loop.

**Primary outcome.** Number of correct responses (N=5280, Mean=7.77, STD=0.48, Median=8.0, IQR=0.0, Range=6.0-8.0).

**Secondary outcomes.** Number of incorrect responses. Mean RT during correct responses.

### 2. **Overt speech marking guidelines**

##### 2.1. Naming Task Primary Outcome

| **Scoring**  **Guidelines** | Score 2/2 marks | **Marking Criteria** |
| --- | --- | --- |
|  |  | The correct word was spoken. |
|  |  | The target word was spoken with valid accompanied words e.g., **acorn** cup, single **bed, coat** hanger, dromedary **camel**, scary **mask**, badminton **racquet**, parchment **scroll**, oak **tree**, erupting **volcano**, ACME (brand) **whistle.**    However, **“**see**-saw”** would receive 0 marks because a playground see-saw does not resemble a handsaw. |
|  |  | A plausible visual misinterpretation occurred, and the answer was semantically unrelated e.g., “hat” or “lampshade” for /acorn/; “memory stick” or “barbecue” for /harmonica/ |
|  |  | A synonym was given e.g., “boat” for /canoe/, “bungalow” for /house/. |
|  |  | Correct answer was spoken then “self-corrected” with a visually misinterpreted answer for /acorn/ e.g., “acorn…thimble”. |
|  |  | The correct answer is spoken and the participant is cut off (5 code) e.g., “a two hump ca-”. We can presume they would say “camel” for /camel/. |
|  |  | Correct answers given in other languages e.g., “harpa” for /harp/ |
|  |  | A phonologically imperfect non-word answer (scoring 1 point for phonology) clearly intended to be the target answer, was spoken “donoro” for /dominoes/ or “darch” for /dart/.    The word spoken must score 1 for phonology e.g., “diamond” for /dominoes/ and “boom” for /boomerang/  would score 0 for semantic because there contain less than 50% matching-phonemes to the target word. |
|  |  | A phonologically imperfect real-word answer (scoring 1 point for phonology) clearly intended to be the target answer, was spoken e.g., “fennel” for /funnel/, “sail” for /snail/. |
|  |  | A phonologically imperfect real or non-word answer is self-corrected to the wrong answer e.g., (non-word) /camel/ “camet no Egypt” and (real word) /dart/ “dart…needle”. |
|  | Score 1/2 marks | **(Vis-sem)** A visually and semantically similar word was spoken e.g., “hazelnut” for /acorn/, “lyre” for /harp/, “xylophone” for /harmonica/, “nutmeg” for /acorn/, “dice” for /dominoes/ |
|  |  | **(Phon-sem)** A Phonologically and semantically similar word was spoken e.g., “harpsichord” for /harp/ and “horse fish” for seahorse |
|  |  | **(RW)** Related word was spoken e.g., “eruption” for /volcano/, “blow” for /whistle/, these words may occur in **circumlocution** e.g., /boomerang/ “you **throw** these things” |
|  |  | **(B)**  brand name was spoken e.g., “ACME Thunderer” for /whistle/ and “Piccolo” for /harmonica/ |
|  |  | **(SPE)** semantic phonological error was spoken e.g., “xenophone” for ‘xylophone’ for the image of the /harmonica/ |
|  | Score 0/2 marks | A semantically unrelated and visually unrelated word was spoken e.g., “mushroom” for /boomerang/, “telephone” for/volcano/ |
|  |  | The answer spoken was clearly intended to be the correct word but scored 0 for phonology e.g., “erm…marker” for /harmonica/. |

##### 2.2. Repetition Task Primary Outcome

| **Scoring**  **Guidelines** | Score 2/2 marks | The target answer and nothing else was spoken with 100% correct phonemes in the correct order. |
| --- | --- | --- |
|  | Score 1/2 marks | The answer was spoken with 50% or more correct phonemes between target and error word.  An answer other than the target word may not score more than 1 mark (this differs from Naming where maximum marks can be achieved through visual misinterpretation). |
|  |  | The answer spoken was a non-word e.g., “sider” for /spider/ , “frotel” for /hotel/, “sudent” for /student/. |
|  |  | The word spoken encompasses the target word e.g. “attribute” for /tribute/  or “remarriage” for /marriage/ |
|  | Score 0/2 marks | The answer was spoken with <50% matching phonemes between target and error word, also known as a neologism. |

##### 2.3. Reading Task Primary Outcome

| **Scoring**  **Guidelines** | Score 2/2 marks | The target answer and nothing else was spoken with 100% correct phonemes in the correct order. |
| --- | --- | --- |
|  | Score 1/2 marks | The answer was spoken with 50% or more correct phonemes between target and error word e.g., “lightning” for /listening/.  An answer other than the target word may not score more than 1 mark (this differs from Naming where maximum marks can be achieved through visual misinterpretation). |
|  |  | The answer spoken was a non-word e.g., “lissing” for /listening/ (reading) |
|  | Score 0/2 marks | The answer was spoken with <50% matching phonemes between target and error word. |
|  |  | The person spells out the word e.g., /to/ T-O or /dwelb/ D-W-E-L-B. This demonstrates spelling rather than blending sounds together for reading aloud. |

##### 2.4. General guidelines across tasks

| **Scoring guidelines** | 0/0 marks | No words are spoken by the speaker. |
| --- | --- | --- |
|  |  | Recording only contains unintelligible words. |
|  | Comments | Allophones of standard English were accepted throughout to mark inclusively of regional British accents e.g., Scottish, Welsh, Nigerian, Indian and Spanish.  For example, in the Reading task “Lis-nin’” for /listening/ is acceptable. |

##### 2.5. Speech data annotation and extraction of performance measures

The optional speech production tasks were manually analysed offline by 6 trained expert raters with high inter-rater reliability (average intraclass correlation=0.86). Inconsistencies amongst raters were resolved by a certified speech and language therapist. Given the labour-intensive nature of scoring these speech production tasks, we discuss normative ranges on single-word speech production tasks only (Reading, Repetition and Naming) derived from 130 control participants. Quantitative speech metrics from the overt picture description task (requiring spontaneous speech production of full sentences in discourse) will be analysed and compiled as part of a separate paper, dedicated to language performance of this patient cohort.

The primary outcome measures for the speech tasks were ‘correct’ (all trials with a maximum possible score of 2/2, see below for scoring guidelines) and ‘incorrect (<2/2). Individual speech trials were excluded from analysis if they met any of the following conditions:

1) Contained high level of background noise which precluded reliable transcription

2) Did not contain audio sound in the speech files

3) The length of the speech files was shorter than 200 milliseconds, suggesting that the trial was accidentally skipped.

### 3. **Cognitive and speech data pre-processing**

Given the remote nature of the cognitive testing, in order to ensure that the normative data was derived from fully engaged healthy participants who understood the task instructions, we implemented three levels of data filtering. These stages include filtering at subject-level, task-level, and trial-level as described below.

**Subject-level data filtering**. Exclusion criteria at participant level was based on self-reported (see supplementary methods) presence of:

I. Neurological co-morbidities (N=407).
II. Family history of young-onset (<60 years) dementia (N=91).
III. Lack of engagement in the tasks (N=16).

An additional exclusion criterion was failure on two simple cognitive screens (orientation to time: <2/4 correct and pear cancellation: <80%), (N=6).

**Task-level data filtering.** The distributions of the primary and secondary outcome measures for each of the 22 tasks were examined to identify outlier performance. Considering the short duration of each task (~2-3 minutes), and the unsupervised context of assessment, individual tasks were excluded for each participant based on the following time- and performance-based heuristic criteria:

I. Changing browser tabs or minimising the assessment browser ≥3 times, or for >10 seconds (s) at a time, during each task.
II. >1 minute keyboard inactivity during each task (suggestive of lack of engagement).
III. Failure on practice trials on any given task.
IV. Outlier performance, defined as control group mean ±3 standard deviations (std). The exclusion criteria were less punitive for tasks with low variability and ceiling effect; for instance, in the Orientation task (where 1 point is acquired for correct orientation to time, day, month, and year), using this approach would have excluded any participants with less than the maximum possible score). Thus, a less stringent criteria of a score <2 (%50 correct) was adopted for this task.
V. Failure to make a response on >50% of the trials within the allocated task duration.
VI. For the optional speech production tasks requiring manual annotations (see Supplementary Results 1.2.), speech tasks from controls (N=1) with pathological speech were excluded.

**Trial-level data filtering.** The trial-by-trial variability in performance within each task was examined for signs of non-engagement. This approach was task-dependent, as non-engagement manifested differently depending on the rules of the task:

I. Where the primary outcome measure of a task was response time-based (e.g., Simple Reaction Time), trials with RT<200 milliseconds were excluded as this was deemed neurobiologically implausible and likely due to premature or repeated motor responses. Tasks where >50% of trials fulfilled this criterion, coupled with unusually low accuracy (defined as <2 std below group mean performance) were excluded.
II. Trials with excessively prolonged RTs were excluded when determining RT (defined as >3 std above the mean RT of the normative sample for that task), as a sign of technical errors or non-engagement.
III. For the optional speech production tasks requiring manual annotations, trials were excluded if there were technical or quality sound recording problems (see Supplementary Results 2.5.).

### 4. **Bayesian predictive modelling**

#### 4.1. Description of Bayesian models

**State-of-the-art Bayesian posterior predictions for modelling the relationship between cognition and confounding factors in controls**. Bayesian regression analyses containing all 8 covariates were performed separately for all tasks, to estimate the effects of each of the covariates on individual task performance. For the optional speech production tasks (repetition, naming, reading) with smaller normative sample (N=130), it was not possible to include depression, anxiety and dyslexia as covariates.

Bayesian linear regression was employed in tasks where residuals followed a normal distribution, otherwise, aggregated binomial Bayesian regression were performed. Sampling from the posterior was executed using the Markov Chain Monte Carlo (MCMC) algorithm; four parallel chains with 5000 samples each were employed. The fit of the model was assessed via examination of chain convergence, sampling efficiency, chain autocorrelation, geometric ergodicity, and via posterior/prior predictive checks. The covariates were normalised around zero if continuous (e.g., age), otherwise binarized. In models with a normal likelihood function, the primary cognitive outcome measure was additionally standardised. Weakly informative standard priors were employed throughout. P-values were computed using standard Region of Practical Equivalence (ROPE) metrics, with null ranges of ±0.1 std for linear models and ±p /√3 log probability for binomial models.^9^

Below you can find the equations used for conducting the Bayesian models.

#### 4.2. Mathematical form of the Bayesian models

##### 4.2.1. Normative model with normal likelihood function

The model is applied to the following cognitive tasks with normally distributed residuals in the primary outcome measure derived from controls: Paired Associates Learning, Digit Span, Spatial Span, Blocks, Odd One Out, Simple Reaction Time.

$${Cognition}_{i} \sim Normal\left( \mu_{i},\sigma\right)$$

$$\mu_{i}=\alpha+\beta_{age}{Age}_{i}+\beta_{sex}{Sex}_{i}+ \beta_{education\_Alevels}{education\_Alevels}_{i}+ \beta_{education\_bachelors}{education\_bachelors}_{i}+ \beta_{education\_postBachelors}{education\_postBachelors}_{i}+ \beta_{device\_phone}{device\_phone}_{i}+ \beta_{device\_tablet}{device\_tablet}_{i}+ \beta_{english\_secondLanguage}{english\_secondLanguage}_{i}+ \beta_{depression}{depression}_{i}+ \beta_{anxiety}{anxiety}_{i}+ \beta_{dyslexia}{dyslexia}_{i}$$

$$\alpha\sim Normal(0,1.5)$$

$$\beta_{age} \sim Normal(0,1.5)$$

$$\beta_{sex} \sim Normal(0,1.5)$$

$$\beta_{education\_Alevels} \sim Normal(0,1.5)$$

$$\beta_{education\_bachelors} \sim Normal(0,1.5)$$

$$\beta_{education\_postBachelors} \sim Normal(0,1.5)$$

$$\beta_{device\_phone} \sim Normal(0,1.5)$$

$$\beta_{device\_tablet} \sim Normal(0,1.5)$$

$$\beta_{english\_secondLanguage} \sim Normal(0,1.5)$$

$$\beta_{depression} \sim Normal(0,1.5)$$

$$\beta_{anxiety} \sim Normal(0,1.5)$$

$$\beta_{dyslexia} \sim Normal(0,1.5)$$

$$\sigma\sim Exp(1)$$

Where $\mu_{i}$ represents the estimated performance for a given participant, and $\sigma$ represents the standard deviation of the model. $\beta$ stands for the beta coefficients and its scale is in standard deviation units. Priors are defined using normal distributions with a weakly informative mean and standard deviation. Note, the dependent variable is z-scored.

##### 4.2.2. Normative model with binomial likelihood function

The model is applied to the following cognitive tasks with non-gaussian distribution of residuals in the primary outcome measure derived from controls: Orientation, Task Recall, Language Comprehension, Semantic Judgement, Trail-making, Rule Learning, Pear Cancellation, Auditory Attention, Choice Reaction Time, Motor Control, Gesture Recognition, and Calculation.

$${Cognition}_{i} \sim Binomial\left( N_{i},p_{i} \right)$$

$${logit(p}_{i})=\alpha+\beta_{age}{Age}_{i}+\beta_{sex}{Sex}_{i}+ \beta_{education\_Alevels}{education\_Alevels}_{i}+ \beta_{education\_bachelors}{education\_bachelors}_{i}+ \beta_{education\_postBachelors}{education\_postBachelors}_{i}+ \beta_{device\_phone}{device\_phone}_{i}+ \beta_{device\_tablet}{device\_tablet}_{i}+ \beta_{english\_secondLanguage}{english\_secondLanguage}_{i}+ \beta_{depression}{depression}_{i}+ \beta_{anxiety}{anxiety}_{i}+ \beta_{dyslexia}{dyslexia}_{i}$$

$$\alpha\sim Normal(0.5,1)$$

$$\beta_{age} \sim Normal(0,1)$$

$$\beta_{sex} \sim Normal(0,1)$$

$$\beta_{education\_Alevels} \sim Normal(0,1)$$

$$\beta_{education\_bachelors} \sim Normal(0,1)$$

$$\beta_{education\_postBachelors} \sim Normal(0,1)$$

$$\beta_{device\_phone} \sim Normal(0,1)$$

$$\beta_{device\_tablet} \sim Normal(0,1)$$

$$\beta_{english\_secondLanguage} \sim Normal(0,1)$$

$$\beta_{depression} \sim Normal(0,1)$$

$$\beta_{anxiety} \sim Normal(0,1)$$

$$\beta_{dyslexia} \sim Normal(0,1)$$

Where $N_{i}$ represents the total number of trials available for a given participant on a specific task, and $p_{i}$ represents the probability of success for a given participant. $\beta$ stands for the beta coefficients and its scale is in log-odd units. Priors are defined via normal distributions with a weakly informative mean and standard deviation. Note, the dependent variable is not z-scored and is in original units.

The code used to run all of the Bayesian regression models is made publicly available upon publication.

### 5. **Results**

##### 5.1. Descriptive statistics and analysis of normative sample.

**Supplementary Table 1.** Demographic characteristics of the normative (N=6364) and stroke (N=90) samples

|  | **Neurologically Healthy Sample** (N=6364) | **Stroke Survivors Sample**  (N=90) |
| --- | --- | --- |
| **^** Age in years  (Mean±SD; Range) | 60.8±10.1; 40-95 | 62.1±13.9; 26-90 |
| **^** Biological sex (%Male:Female) | 42.70:57.30 | 71.1: 28.9 |
| **^ Education %:**  Secondary school ≤16 years  Secondary school aged 18 years  Bachelor’s degree  Postgraduate Degree | 15.5 (N=984)  37.2 (N=2364)  28.8 (N=1835)  18.5 (N=1181) | 12.2 (N=11)  47.8 (N=43)  20.0 (N=18)  20.0 (N=18) |
| **^ Device used %:**  Smart Phone  Tablet  Computer | 20.5 (N=1307)  20.8 (N=1325)  58.7 (N=3732) | 4.4 (N=4)  87.8 (N=79)  7.8 (N=7) |
| **^** With English as second language %: | 3.6 (N=231) | 33.7 (N=31) |
| **^ †** % with Anxiety Diagnosis | 5.4 (N=343) | 1.1 (N=1) |
| **^ †** % with Depression Diagnosis | 7.3 (N=466) | 3.3 (N=3) |
| **^ †** % with Dyslexia Diagnosis | 4.0 (N=255) | 0.0 (N=0) |
| **% with Cerebrovascular disease risk factors**  Ischemic heart disease  Diabetes  High blood pressure  High cholesterol  Kidney disease  Alcohol Dependency  Over-weight  Long-term smoker  Ex-smoker | 4.2 (N=270)  6.6 (N=416)  27.1 (N=1724)  20.1 (N=1282)  1.4 (N=87)  1.2 (N=78)  29.7 (N=1889)  3.9 (N=253)  24.5 (N=1559) | 13.04 (N = 12)  17.39 (N=16)  42.39 (N=39)  19.57 (N=18)  1.09 (N=1)  1.09 (N=1)  13.04 (N=12)  4.35 (N=4)  18.48 (N=17) |
| **Time (minutes) taken to complete the assessment** (Median±IQR; Range)  Full IC3 battery, excluding participant-initiated breaks | 38.4±7.5; 25.15-196.94 | 55.92±27.42; 36.93-187.09 |
| **Time (minutes) taken to complete the questionnaires**  (Median±IQR; Range) | 3.01±0.98; 1.36-28.84 | 4.94±3.42; 1.75-19.59 |
| **Stroke representation** | | |
| Days since stroke  (Median± IQR; Range) |  | 5.0±31.0; 0-556 |
| **% Stroke Phase:**  Acute (0-14 days)  Subacute (15-180)  Chronic phase (>180 days) |  | 67.8 (N=61)  18.9 (N=17)  13.3 (N=12) |
| **% Stroke Aetiology:**  Ischaemic  Haemorrhagic |  | 92.2 (N=83)  7.8 (N=7) |
| **% Hemisphere affected**  Left  Right  Bilateral |  | 50.0 (N=45)  40.0 (N=36)  10.87 (N=9) |
| MOCA (Mean±SD; Range) |  | 21.91±5.95; 3-30 |
| NIHSS (Mean±SD; Range) |  | 3.03±2.35; 0-9 |
| Note. ‘†’= Self-reported diagnosis with concurrent symptoms and receiving anxiolytic / antidepressants medication at the time of the assessment. ^= Covariates that were accounted for in the normative sample. MOCA=Montreal Cognitive Assessment. NIHSS= National Institute of Health Stroke Scale. | | |

##### 5.2. Supplementary Figure 2. Lesion overlap map.

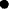

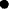

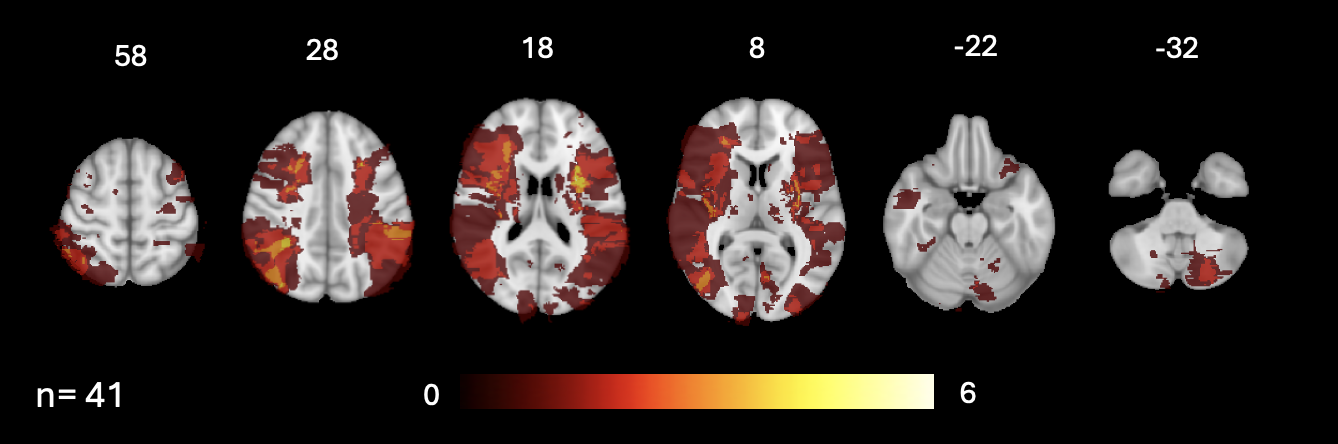

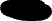

*Supplementary Figure 2. Lesion overlap map at 3-months post-stroke in subset of N=41 patients that underwent research MRI as part of the IC3 study.*

##### 5.3. Internal consistency

We used the well-established method of split-half reliability to assess internal consistency. In this method, trials in each task are randomly divided into two parts, and a score is computed for each half. Subsequently, a two-part reliability coefficient, such as the Spearman-Brown adjusted Pearson correlation, is calculated between the scores from the two halves. Since split-half reliability is assessed based on aggregates of trials, it circumvents practical issues associated with models for individual trials. One potential factor that could influence the accuracy of split-half reliability estimates is the method used to split the task trials. To ensure the generalisability of our findings, the splitting was done via 5000 random permutations, creating a distribution of split-half estimates across wide range of splits. Here, we report the median split-half internal reliability coefficient across these iterations, which was conducted on the large normative sample (N=6364).

##### 5.4. Test-retest reliability

Test-retest reliability. 94 older adults (sub-cohort A in Figure 1) completed the IC3 twice (median gap=30, IQR=100 days). Consistency between sessions was assessed via well-established metrics: (a) group difference analyses, (b) equivalence testing, and (c) correlation analyses. Parametric and non-parametric statistical tests were employed, conditional on the distributions of the residuals.

**Supplementary Table 2.** Internal consistency (full normative cohort) and test-retest reliability (sub-cohort A) analyses of the older adult controls.

|  |  | **Internal Consistency** | | **Test-retest reliability sub-study** | | | | | | | |
| --- | --- | --- | --- | --- | --- | --- | --- | --- | --- | --- | --- |
| **Domain** | **Task** | **N** | **Alpha** | **N** | **Mean Time 1** | **Mean Time 2** | **Group mean difference P-value** | **Effect size** | **Test of equivalence P-value** | **R-value** | **Normally Distributed** |
| **Memory** | Orientation | 6290 | 0.14† | 92 | 3.88 | 3.9 | 0.74 | 0.07 | <0.001 | 0.1† | No |
|  | Task Recall | 4782 | 0.04† | 91 | 3.79 | 3.87 | 0.34 | 0.2 | <0.001 | 0.31† | No |
|  | Paired Associates Learning | 5721 | 0.73 | 87 | 4.68 | 4.97 | 0.29 | 0.22 | 0.03 | 0.31 | Yes |
|  | Digits Span | 5279 | 0.73 | 86 | 6.69 | 7.0 | 0.1 | 0.24 | 0.03 | 0.59 | Yes |
|  | Spatial Span | 5867 | 0.70 | 90 | 5.08 | 5.22 | 0.46 | 0.13 | 0.01 | 0.45 | Yes |
| **Language** | Comprehension | 5025 | 0.76 | 91 | 18.44 | 18.52 | 0.49 | 0.05 | 0.01 | 0.45 | No |
|  | Semantic Judgement | 4995 | 0.94 | 92 | 22.4 | 22.58 | 0.46 | 0.14 | <0.001 | 0.62 | No |
|  | Naming | 132ˆ | 0.79 | 85 | 0.97 | 0.98 | 0.41 | 0.13 | <0.001 | 0.46 | No |
|  | Reading | 132ˆ | 0.72 | 87 | 0.99 | 0.99 | 0.19 | 0.23 | <0.001 | 0.12† | No |
|  | Repetition | 132ˆ | 0.83 | 86 | 0.99 | 0.98 | 0.12 | 0.33 | <0.001 | -0.05† | No |
| **Executive** | Blocks | 5039 | 0.70 | 54^‡^ | 6.52 | 7.28 | 0.1 | 0.22 | 0.03 | 0.77 | Yes |
|  | Trail-making* | 5189 | 0.81 | 82 | 1.02 | 1.05 | 0.92 | 0.01 | <0.001 | 0.08† | No |
|  | Odd One out | 5672 | 0.75 | 91 | 10.73 | 11.23 | 0.1 | 0.23 | 0.03 | 0.59 | Yes |
|  | Rule Learning | 5485 | 0.83 | 85 | 6.75 | 7.24 | 0.1 | 0.27 | 0.04 | 0.42 | No |
| **Attention** | Pear Cancellation | 6219 | 0.88 | 85 | 0.98 | 0.99 | 0.1 | 0.41 | <0.001 | 0.3† | No |
|  | Simple Reaction Time* | 5510 | 0.95 | 91 | 390.66 | 386.12 | 0.66 | 0.06 | <0.001 | 0.68 | Yes |
|  | Auditory Attention | 5757 | 0.73 | 91 | 35.24 | 35.44 | 0.35 | 0.15 | <0.001 | 0.32† | No |
| **Motor Ability** | Choice Reaction Time | 5181 | 0.88 | 87 | 54.59 | 53.9 | 0.65 | 0.08 | 0.01 | 0.28 | Yes |
|  | Motor Control | 5208 | 0.88 | 93 | 28.96 | 28.95 | 0.97 | 0.01 | <0.001 | 0.14† | No |
| **Numeracy** | Calculation | 5681 | 0.33† | 91 | 7.86 | 7.88 | 0.82 | 0.04 | <0.001 | 0.09† | No |
| **Praxis** | Gesture Recognition | 5280 | 0.05† | 91 | 7.48 | 7.55 | 0.59 | 0.09 | <0.001 | 0.36† | No |
| Note: ‘N’= Sample size. ‘ˆ’= Lower sample size due to tasks requiring manual transcription of overt speech. ‘Alpha’= 5000 iterations of permutation-based randomly split-half sampled trial-level data were used to calculate internal consistency. ‘†’= Unreliable alpha values due to lack of variance in controls and low number of trials. Nevertheless, test-retest reliability suggests that all these tasks are stable across time. ‘Group Difference P-value’= Obtained from a paired-samples t-test or Wilcoxon test depending on the stated distribution. ‘Effect size’= Cohen’s D. ‘Test of equivalence P-value’= P-value obtained from a parametric on non-parametric test of equivalence dependent on the stated distribution; significant p-values equate equivalence. P-values are FDR-corrected. ‘R-value’= Coefficient obtained from a Pearson or Spearman correlation dependent on the stated distribution. ‘*’= Indicates tasks where higher values suggest worse performance. ‘‡’= Lower N caused by exclusion of 32 participants whose version of Blocks differed between sessions due to improvements in task design. ‘†’ in R-value = Indicates that the correlation values may be low due to lack of variance in both sessions, despite highly similar mean values between sessions. | | | | | | | | | | | |

##### 5.5. Diurnal effects

The effect of inter-session time interval and that of time of day when the assessment was performed was evaluated using multi-level random-intercept Bayesian regression models. The main predictor of interest were the interaction terms: session*time of day, and session*inter-session time interval in days, whilst accounting for the main effects of session, age, gender, education, device, time of day and inter-session interval. No significant diurnal effects were found (in interaction terms) suggesting that time related factors do not modulate change in cognitive ability across the two sessions (see Supplementary Table 2). Furthermore, there was no significant main effect of session across all tasks (p<0.27), reconfirming the test-retest reliability between sessions and their independence from diurnal factors and demographic variables.

**Supplementary Table 3**. Multi-level Bayesian analyses showing the effect of inter-session time interval and time of day when the assessments were performed on across two testing timepoints (sub-cohort A).

| **Domain** | **Task** | **N** | **Session * Time in Days** | **95% Credible Interval Days** | **Session * Time in Hours** | **95% Credible Interval Hours** | **Normally Distributed** |
| --- | --- | --- | --- | --- | --- | --- | --- |
| **Memory** | Orientation | 92 | 0.54 | [-0.25, 1.55] | 0.54 | [-0.14, 0.17] | No |
|  | Task Recall | 91 | 0.61 | [-1.11, 0.55] | 0.61 | [-0.05, 0.22] | No |
|  | Paired Associates Learning | 87 | 0.87 | [-0.24, 0.22] | 0.87 | [-0.4, 0.07] | Yes |
|  | Digits Span | 86 | 0.73 | [-0.22, 0.22] | 0.73 | [-0.12, 0.39] | Yes |
|  | Spatial Span | 90 | 0.54 | [-0.48, 0.0] | 0.54 | [-0.3, 0.23] | Yes |
| **Language** | Comprehension | 91 | 0.77 | [-0.22, 0.35] | 0.77 | [-0.06, 0.09] | No |
|  | Semantic Judgement | 92 | 0.62 | [-0.28, 0.67] | 0.62 | [-0.19, 0.03] | No |
|  | Naming | 85 | 0.62 | [-0.22, 0.54] | 0.62 | [-0.13, 0.05] | No |
|  | Reading | 87 | 0.54 | [-1.83, 0.28] | 0.54 | [-0.24, 0.07] | No |
|  | Repetition | 86 | 0.62 | [-0.44, 0.75] | 0.62 | [-0.2, 0.09] | No |
| **Executive** | Blocks | 54^‡^ | 0.62 | [-0.17, 0.31] | 0.62 | [-0.29, 0.14] | Yes |
|  | Trail-making* | 82 | 0.54 | [-1.87, 0.11] | 0.54 | [-0.07, 0.29] | No |
|  | Odd One out | 91 | 0.61 | [-0.35, 0.07] | 0.61 | [-0.1, 0.38] | Yes |
|  | Rule Learning | 85 | 0.77 | [-0.36, 0.23] | 0.77 | [-0.1, 0.54] | No |
| **Attention** | Pear Cancellation | 85 | 0.61 | [-0.86, 0.55] | 0.61 | [0.04, 0.36] | No |
|  | Simple Reaction Time* | 91 | 0.62 | [-0.3, 0.08] | 0.62 | [-0.13, 0.31] | Yes |
|  | Auditory Attention | 91 | 0.61 | [-0.23, 0.8] | 0.61 | [-0.09, 0.1] | No |
| **Motor Ability** | Choice Reaction Time* | 87 | 0.54 | [0.01, 0.45] | 0.54 | [-0.21, 0.27] | No |
|  | Motor Control | 93 | 0.61 | [-0.11, 0.66] | 0.61 | [-0.08, 0.14] | No |
| **Numeracy** | Calculation | 91 | 0.61 | [-1.15, 0.62] | 0.61 | [-0.13, 0.16] | No |
| **Praxis** | Gesture Recognition | 91 | 0.61 | [-0.18, 0.81] | 0.61 | [-0.08, 0.13] | No |
| Note: ‘N’ = Sample size after pre-processing. ‘Session * Time in Days’ = P-value for the interaction between the session number and the time difference in days between sessions for each cognitive task. ‘Session * Time in Hours’ = P-value for the interaction between the session number and the time difference in hours between sessions for each cognitive task. ‘‡’ = Lower N caused by exclusion of 32 participants whose version of Blocks differed between sessions due to user-suggested improvements in task design. | | | | | | | |

##### 5.6. Effects of unsupervised testing environment

The effect of unsupervised testing environment was assessed by comparing results from supervised (in-person) and unsupervised (remote) administration of IC3 in a subset of 44 older control adults (sub-cohort B) who completed the assessment twice (median inter-session interval=100 days; IQR=89.92). The order in which the assessments were administered were counterbalanced to minimise potential order effects. The testing environment effects were assessed using group difference analyses and equivalence testing (see Supplementary Table 3).

Overall, we found little to no effect of environment in the cognitive battery, except for two tasks that measure hand motor dexterity (e.g., Choice Reaction Time and Motor Control). Namely, we found that motor ability was better within a supervised setting, than in a remote setting (Supplementary Table 3). Whilst performance on these tasks is strongly dependent on the type of device that the participant uses (as seen in Figure 3), we posited that the group differences may be explained by changes in device between remote and supervised settings. To test this, we conducted two random-intercept Bayesian regression models, with participant ID as the grouping factor, and with age, sex, education, device and environment as independent variables. Posterior predictions indicated no effect of environment after accounting for device in either the Choice Reaction Time (un-corrected p-value = .36, 95% CI = [-.44, .26]) or the Motor Control Tasks (un-corrected p-value = .28, 95% CI = [-.36,.60]). However, we did find a strong effect of device in both tasks (un-corrected p = .001, 95% CI = [.33,1.16], and p = .0002, 95% CI = [.58,1.87], respectively).

In the Choice Reaction Time, which captures how quickly one can switch between selecting the left or right side of the screen, we found that using a touch screen device leads to quicker responses, compared to a mouse, since less dexterity is needed to make the hand movement. For the motor control task, which measures how accurately participants can hit a small moving target, having a mouse pointer led to better accuracy as this improved the resolution of the spatial localisation of the responses, compared to a touchscreen. These effects were consistent with those presented in Figure 3 (main manuscript). Overall, we demonstrate that the IC3 provides a reliable and comparable measure of cognition in both remote and supervised settings.

**Supplementary Table 4**. Effects of supervised versus unsupervised testing environment on cognition in sub-cohort B.

| **Domain** | **Task** | **N** | **Mean Remote** | **Mean Supervised** | **P-value** | **Effect size** | **Test of equivalence** | **Normally Distributed** |
| --- | --- | --- | --- | --- | --- | --- | --- | --- |
| **Memory** | Orientation | 43 | 3.98 | 4.0 | 1.0 | 0.22 | <0.001 | No |
|  | Task Recall | 43 | 3.91 | 3.91 | 1.0 | 0.0 | <0.001 | Yes |
|  | Paired Associates Learning | 41 | 4.95 | 4.88 | 1.0 | 0.06 | 0.01 | Yes |
|  | Digits Span | 42 | 7.05 | 6.76 | 0.63 | 0.24 | 0.03 | Yes |
|  | Spatial Span | 43 | 5.12 | 5.4 | 0.63 | 0.27 | 0.03 | Yes |
| **Language** | Comprehension | 44 | 18.91 | 19.05 | 1.0 | 0.11 | 0.02 | No |
|  | Semantic Judgement | 43 | 22.74 | 22.63 | 0.63 | 0.14 | <0.001 | No |
|  | Naming | 44 | 0.98 | 0.97 | 0.86 | 0.11 | 0.01 | No |
|  | Reading | 43 | 1.0 | 1.0 | 1.0 | 0.0 | <0.001 | No |
|  | Repetition | 42 | 0.99 | 0.99 | 0.75 | 0.26 | <0.001 | No |
| **Executive** | Blocks | 39 | 7.72 | 7.28 | 0.88 | 0.14 | 0.02 | Yes |
|  | Trail-making* | 40 | 0.32 | 1.48 | 0.07 | 0.57 | 0.03 | No |
|  | Odd One out | 41 | 10.93 | 11.32 | 0.63 | 0.21 | 0.02 | Yes |
|  | Rule Learning | 40 | 7.0 | 7.05 | 1.0 | 0.03 | 0.02 | No |
| **Attention** | Pear Cancellation | 42 | 0.99 | 0.99 | 1.0 | 0.05 | <0.001 | No |
|  | Simple Reaction Time* | 42 | 379.6 | 393.48 | 0.72 | 0.19 | 0.02 | Yes |
|  | Auditory Attention | 43 | 35.58 | 35.51 | 1.0 | 0.08 | <0.001 | No |
| **Motor Ability** | Choice Reaction Time | 43 | 51.35 | 59.4 | **<0.001** | 1.11 | **0.05** | No |
|  | Motor Control | 44 | 29.41 | 28.09 | **<0.001** | 0.69 | **0.05** | No |
| **Numeracy** | Calculation | 43 | 7.91 | 7.95 | 1.0 | 0.13 | <0.001 | No |
| **Praxis** | Gesture Recognition | 43 | 7.72 | 7.56 | 0.4 | 0.31 | 0.01 | No |
| Note: ‘N’ = Sample size after preprocessing. ‘P-value’ = Obtained from a paired-samples t-test when the sample is normally distributed or from a Wilcoxon test when the sample is not. ‘Effect size’ = Cohen’s D. ‘Test of equivalence’ = P-value obtained from a parametric test of equivalence when the sample is normally distributed, or from a non-parametric test if the sample is not. Here significant p-values >0.05 equate equivalence. All p-values are FDR-corrected and shown in bold if significant. ‘*’ = Indicates tasks where higher values suggest worse performance. | | | | | | | | |

##### 5.7. Learning effects

To examine the potential effect of learning on repeated testing, 30 older control participants (Sub Cohort C) completed the IC3 four times over the course of two weeks. To examine the change in performance over the four sessions, random-intercept multi-level Bayesian regression models were performed separately for each task, accounting for age, gender, education, and device. Overall, the results indicate little to no effect of learning across tasks (Supplementary Table 4); Out of 22 tasks, only one task (Odd One Out) showed significant learning effect between the 1^st^ and 4^th^ timepoint. Nevertheless, there was no learning effect for this task between the first two sessions (*P*=.08 un-corrected).

This task examines high-level reasoning coupled with visuo-spatial pattern processing with increasing difficulty over a period of two minutes. Given the nature of the task, it is probable that participants became quicker at identifying patterns after playing the task 4 times in quick succession, leading to higher number of patterns identified overall in the allotted time. Therefore, except for the Odd One Out task, all tasks in the IC3, can be deployed up to 4 times in a span of two weeks, with minimal expected learning effect. For the Odd One Out task, it can be safely deployed twice in the span of a week, followed by a longer break to minimise learning effects. In reality, participants will unlikely be required to complete the assessment at such a high frequency and short time interval. Thus, IC3 assessment has minimal learning effects within the expected time interval of cognitive testing.

**Supplementary Table 5.** Learning effects analyses in a sample of older adults that completed 4 consecutive IC3 sessions in the span of two weeks (sub-cohort C)

| **Domain** | **Task** | **N** | **Mean T1** | **Mean T4** | **Beta** | **P-value** | **95% Credible Interval** | **Normally Distributed** |
| --- | --- | --- | --- | --- | --- | --- | --- | --- |
| **Memory** | Orientation | 29 | 3.9 | 3.97 | 0.62 | 0.3 | [-0.91, 2.21] | No |
|  | Task Recall | 29 | 3.72 | 3.79 | 0.27 | 0.33 | [-0.79, 1.33] | Yes |
|  | PAL | 28 | 4.5 | 5.04 | 0.4 | 0.22 | [-0.05, 0.84] | Yes |
|  | Digits Span | 30 | 6.43 | 7.07 | 0.39 | 0.22 | [-0.01, 0.79] | Yes |
|  | Spatial Span | 26 | 4.85 | 5.12 | 0.28 | 0.3 | [-0.14, 0.7] | Yes |
| **Language** | Comprehension | 28 | 17.89 | 18.64 | 0.48 | 0.22 | [0.06, 0.91] | No |
|  | Semantic Judgement | 29 | 22.45 | 22.59 | 0.29 | 0.35 | [-0.45, 1.06] | No |
|  | Naming | 25 | 0.96 | 0.98 | 0.62 | 0.22 | [0, 1.27] | No |
|  | Reading | 26 | 0.99 | 0.99 | 0.02 | 0.33 | [-1.21, 1.25] | No |
|  | Repetition | 27 | 1.05 | 1.06 | 0.05 | 0.33 | [-1.41, 1.5] | No |
| **Executive** | Blocks | 25 | 8.08 | 9 | 0.35 | 0.22 | [0.03, 0.66] | Yes |
|  | Trail-making* | 26 | 0.58 | 0.5 | -0.26 | 0.35 | [-1.04, 0.52] | No |
|  | Odd One out | 30 | 10.37 | 12.03 | 0.76 | **<.001** | [0.45, 1.08] | Yes |
|  | Rule Learning | 28 | 6.79 | 7.68 | 0.66 | 0.15 | [0.19, 1.13] | No |
| **Attention** | Pear Cancellation | 28 | 0.98 | 1 | 1.76 | 0.06 | [0.47, 3.25] | No |
|  | SRT* | 28 | 413.87 | 391.75 | -0.2 | 0.3 | [-0.4, 0.0] | Yes |
|  | Auditory Attention | 29 | 35.31 | 35.1 | -0.28 | 0.35 | [-0.87, 0.3] | No |
| **Motor Ability** | CRT | 30 | 46.87 | 48.27 | 0.11 | 0.45 | [-0.08, 0.29] | No |
|  | Motor Control | 30 | 29.13 | 28.7 | -0.19 | 0.41 | [-0.74, 0.36] | No |
| **Numeracy** | Calculation | 28 | 7.79 | 7.86 | 0.29 | 0.33 | [-0.86, 1.46] | No |
| **Praxis** | Gesture Recognition | 29 | 7.55 | 7.59 | 0.09 | 0.36 | [-0.7, 0.9] | No |
| Note: ‘N’ = Sample size after pre-processing. ‘Mean T1’ = Raw scores at Timepoint 1. ‘Beta’ = Refers to the standardised beta coefficient in tasks with normally distributed residuals and to log-odds in those with non-normal distribution. ‘P-value’ = Estimated effect of learning obtained from a Bayesian regression model which accounts for age, gender, education and device. P-values are FDR corrected and shown in bold if significant. ‘*’ = Indicates tasks where higher values suggest worse performance (as measured by raw scores). | | | | | | | | |

##### 5.8. Group differences between healthy controls and patients with stroke

**Supplementary Table 6.** Group differences between 90 patients with stroke and the large normative cohort. The mean group performance for each task is represented in standard deviation units from the controls (deviation from expected units) accounting for 8 clinical and demographic variables. Negative mean values signify that patients performed worse than demographically-matched controls (in SD units). P-values are FDR corrected and are all significant.

| **Domain** | **Task** | **Mean** | **STD** | **P-value** | **Effect size (Cohen’s D)** |
| --- | --- | --- | --- | --- | --- |
| **Memory** | **Orientation** | -1.36 | 2.62 | <0.001 | 0.52 |
|  | **Task Recall** | -1.61 | 2.49 | <0.001 | 0.64 |
|  | **Paired Associates Learning** | -1.35 | 1.13 | <0.001 | 1.19 |
|  | **Digit Span** | -1.76 | 1.4 | <0.001 | 1.26 |
|  | **Spatial Span** | -1.22 | 1.32 | <0.001 | 0.92 |
| **Language** | **Language Comprehension** | -2.54 | 2.63 | <0.001 | 0.97 |
|  | **Semantic Judgement** | -5.07 | 7.53 | <0.001 | 0.67 |
|  | **Naming** | -1.61 | 3.32 | 0.009 | 0.48 |
|  | **Reading** | -2.71 | 5.29 | 0.007 | 0.51 |
|  | **Repetition** | -1.12 | 2.63 | 0.029 | 0.43 |
| **Executive** | **Blocks** | -2.11 | 1.32 | <0.001 | 1.59 |
|  | **Trail-making** | -2.69 | 2.7 | <0.001 | 1.0 |
|  | **Odd One Out** | -1.27 | 1.51 | <0.001 | 0.84 |
|  | **Rule Learning** | -1.62 | 1.56 | <0.001 | 1.04 |
| **Attention** | **Pear Cancellation** | -7.07 | 11.24 | <0.001 | 0.63 |
|  | **Simple Reaction Time** | -2.23 | 1.85 | <0.001 | 1.21 |
|  | **Auditory Attention** | -2.65 | 3.66 | <0.001 | 0.72 |
| **Motor Ability** | **Choice Reaction Time** | -2.54 | 3.95 | <0.001 | 0.64 |
|  | **Motor Control** | -0.92 | 1.83 | <0.001 | 0.5 |
| **Numeracy** | **Graded Calculation** | -3.44 | 4.96 | <0.001 | 0.69 |
| **Praxis** | **Gesture Recognition** | -2.41 | 3.21 | <0.001 | 0.75 |

##### 5.9. Factor analysis

To determine whether the data is suited for factor analysis, the Kaiser–Meyer–Olkin (KMO) test was conducted. The results showed high sampling adequacy, with KMO=0.86. In addition, the Bartlett’s test of Sphericity was conducted to test the null hypothesis that the correlation matrix is an identity matrix. The results rejected the null hypothesis, at p<.001, suggesting the task scores are reasonably correlated with each other and are thus suitable for factor analysis. The optimal number of factors needed for the factor analysis was determined in a data driven way via parallel analysis (i.e. by comparing the factor solution to random data with the same properties as the real data set). The results of the parallel analysis, shown in Supplementary Figure 2, suggested a 6-factor structure.

In light of these results, an exploratory factor analysis (EFA) was conducted, using the psych (v. 2.4.1) package in R, specifying the bi-factor model of cognition: a global factor ‘g’ which loads positively on all test scores, and 6 group factors that each load on subsets of the test scores. All latent factors are orthogonal to one another. Omega function was used, which conducts a factor analysis (with maximum-likelihood estimation) of the dataset, rotates the factors obliquely (using ‘oblimin’ rotation), factors the resulting correlation matrix, followed by a Schmid–Leiman transformation to find general factor loadings. The solution is depicted in Figure 5A (main manuscript). The EFA was shown to be robust (CFI=0.94, RMSEA=0.01) and have good internal consistency (omega=0.79). Factor scores were derived using the ‘tenBerge’, method.

Often the EFA is followed up by a confirmatory factor analysis (CFA). The main difference between EFA and CFA is that in EFA observed task scores are allowed to cross-load freely on several group factors, while in CFA such cross-loadings may be forbidden. For the purpose of deriving the general factor of cognition ‘g’, there is little difference between using CFA and EFA. Subsequently, we wanted to show that an intuitive solution emerges from the data, rather than it being pre-specified. Nevertheless, a CFA was also conducted using the model obtained from the EFA analysis, to confirm there are no meaningful differences between the two. The results indicate a robust fit for the CFA model, CFI=0.91, RMSEA=0.04, and that the two global g factors outputted from the CFA and the EFA are highly correlated, r=0.98.

**Supplementary Figure 3.** Parallel analysis suggesting the presence of six latent factors from the covariance structure of cognitive task scores.

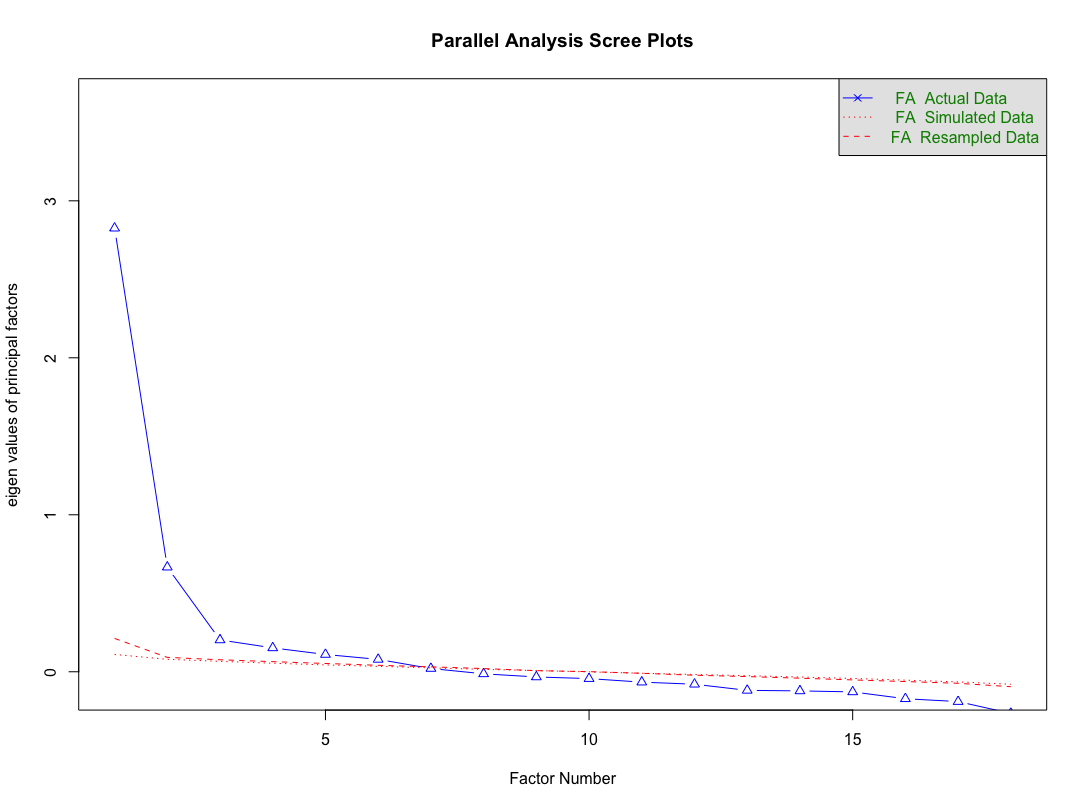

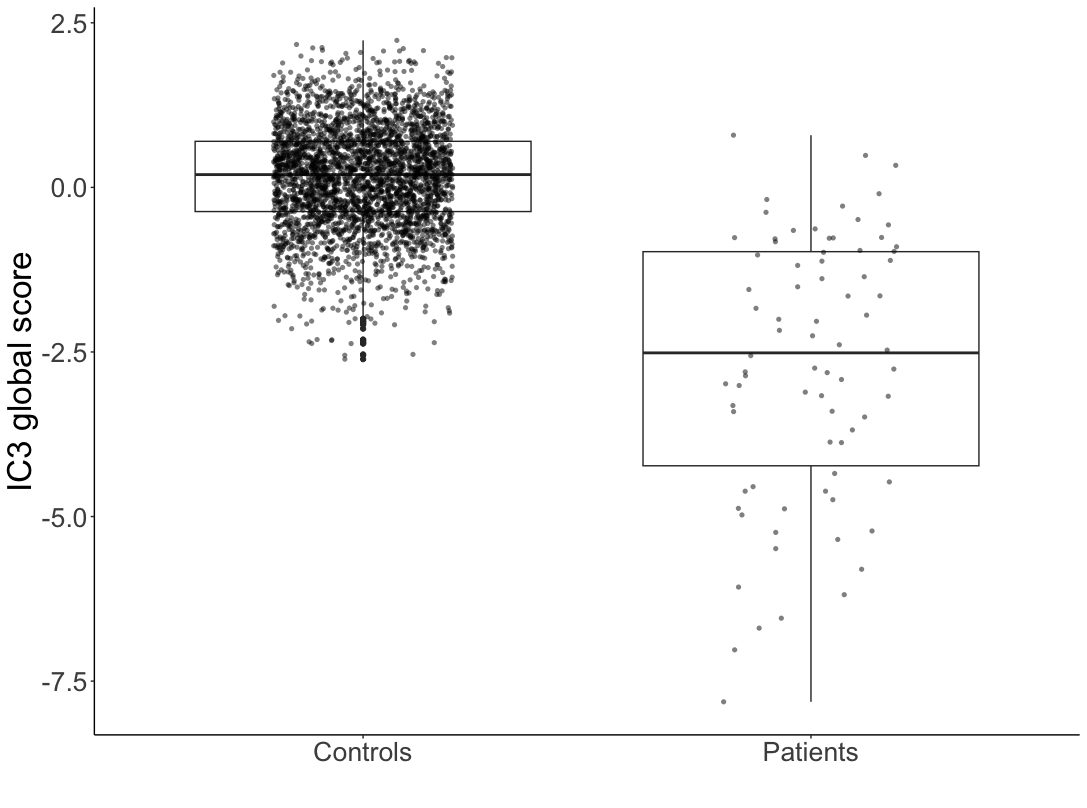

**Supplementary Figure 4.** The IC3 global cognitive score derived from factor analysis shows group differences between patients and controls. The thick horizontal line represents the median, the thin lines represent the IQR.

##### 5.10. Divergent validity

Divergent validity was determined through an anticipated lack of significant correlations between cognitive scores and cholesterol levels at the time of admission. As expected, there were no significant associations (Supplementary Table 5).

**Supplementary Table 7.** Divergent Validity in a cohort of stroke survivors. Cognitive task scores are correlated to admission cholesterol levels. P-values are FDR corrected.

| **Domain** | **Task** | **N** | **R-value** | **P-value** |
| --- | --- | --- | --- | --- |
| **Memory** | **Orientation** | 82 | 0.02 | 0.85 |
|  | **Task Recall** | 67 | -0.13 | 0.41 |
|  | **Paired Associates Learning** | 68 | -0.01 | 0.93 |
|  | **Digit Span** | 70 | 0.09 | 0.57 |
|  | **Spatial Span** | 70 | 0.04 | 0.76 |
| **Language** | **Language Comprehension** | 74 | 0.12 | 0.41 |
|  | **Semantic Judgement** | 74 | 0.12 | 0.41 |
|  | **Naming** | 30 | 0.08 | 0.76 |
|  | **Reading** | 29 | 0.2 | 0.41 |
|  | **Repetition** | 27 | 0.37 | 0.16 |
| **Executive** | **Blocks** | 71 | 0.04 | 0.76 |
|  | **Trail-making** | 82 | -0.1 | 0.5 |
|  | **Odd One Out** | 71 | 0.14 | 0.41 |
|  | **Rule Learning** | 74 | -0.05 | 0.76 |
| **Attention** | **Pear Cancellation** | 79 | 0.09 | 0.57 |
|  | **Simple Reaction Time** | 72 | -0.04 | 0.76 |
|  | **Auditory Attention** | 71 | 0.05 | 0.76 |
| **Motor Ability** | **Choice Reaction Time** | 71 | 0.16 | 0.41 |
|  | **Motor Control** | 71 | 0.07 | 0.71 |
| **Numeracy** | **Graded Calculation** | 68 | 0.09 | 0.57 |
| **Praxis** | **Gesture Recognition** | 68 | -0.03 | 0.79 |

##### 5.11.  IC3 sensitivity compared to MOCA

Tasks on the IC3 were validated against specific tasks on the MOCA, wherever a 1-to-1 mapping existed between the two at both task level and pre-specified cognitive domains level.^10^ In the absence of a gold standard for definition of post-stroke cognitive impairment, clinically derived MOCA scores were assigned as the approximate ‘ground truth’ which the IC3 sensitivity was calculated.^11^ Impairment on MOCA was arbitrarily defined as <total max score on a task, with the exception of memory delayed recall where max-1 was used.

### 6.  **References**

1.	Guerrero-Berroa E, Luo X, Schmeidler J, Rapp MA, Dahlman K, Grossman HT, et al. The MMSE orientation for time domain is a strong predictor of subsequent cognitive decline in the elderly. International Journal of Geriatric Psychiatry. 2009;24(12):1429–37.

2.	Blackwell AD, Sahakian BJ, Vesey R, Semple JM, Robbins TW, Hodges JR. Detecting Dementia: Novel Neuropsychological Markers of Preclinical Alzheimer’s Disease. Dementia and Geriatric Cognitive Disorders. 2003 Dec 11;17(1–2):42–8.

3.	Halai AD, De Dios Perez B, Stefaniak JD, Lambon Ralph MA. Comparing short and long batteries to assess deficits and their neural bases in stroke aphasia [Internet]. Neuroscience; 2020 Nov [cited 2024 Mar 17]. Available from: http://biorxiv.org/lookup/doi/10.1101/2020.11.24.395590

4.	Kay J, Lesser R, Coltheart M. Psycholinguistic assessments of language processing in aphasia (PALPA): An introduction. Aphasiology. 1996 Feb 1;10(2):159–80.

5.	Halai AD, De Dios Perez B, Stefaniak JD, Lambon Ralph MA. Efficient and effective assessment of deficits and their neural bases in stroke aphasia. Cortex. 2022 Oct 1;155:333–46.

6.	Jefferies E, Patterson K, Jones RW, Lambon Ralph MA. Comprehension of concrete and abstract words in semantic dementia. Neuropsychology. 2009;23(4):492–9.

7.	Almaghyuli A, Thompson H, Lambon Ralph MA, Jefferies E. Deficits of semantic control produce absent or reverse frequency effects in comprehension: Evidence from neuropsychology and dual task methodology. Neuropsychologia. 2012 Jul 1;50(8):1968–79.

8.	Massa MS, Wang N, Bickerton WL, Demeyere N, Riddoch MJ, Humphreys GW. On the importance of cognitive profiling: A graphical modelling analysis of domain-specific and domain-general deficits after stroke. Cortex. 2015 Oct 1;71:190–204.

9.	Kruschke JK. Rejecting or Accepting Parameter Values in Bayesian Estimation. Advances in Methods and Practices in Psychological Science. 2018 Jun 1;1(2):270–80.

10.	Gruia DC, Trender W, Hellyer P, Banerjee S, Kwan J, Zetterberg H, et al. IC3 protocol: a longitudinal observational study of cognition after stroke using novel digital health technology. BMJ Open. 2023 Nov 1;13(11):e076653.

11.	Quinn TJ, Richard E, Teuschl Y, Gattringer T, Hafdi M, O’Brien JT, et al. European Stroke Organisation and European Academy of Neurology joint guidelines on post-stroke cognitive impairment. European Stroke Journal. 2021 Sep;6(3):I–XXXVIII.
